# Screening, prevention, and management of maternal acute malnutrition and anemia in Ethiopia: evidence from a longitudinal eCohort study

**DOI:** 10.1101/2025.09.10.25335154

**Authors:** Emma Clarke-Deelder, Theodros Getachew, Tefera Taddele, Getachew Tollera, Katherine Wright, Delayehu Bekele, Günther Fink, Margaret E Kruk, Mariia Kuleba, Nandita Perumal, Catherine Arsenault

## Abstract

**Background:** Undernutrition during pregnancy, including both micronutrient deficiencies and chronic energy deficiency, is an important public health concern with negative consequences for maternal, neonatal, and child health, development, and economic outcomes through the life course. Despite widespread efforts to address undernutrition during pregnancy, there is little evidence on the quality of management of maternal undernutrition in high-burden settings. Data on follow-up care for undernutrition over the course of pregnancy is particularly scarce. In this study, we used a novel longitudinal dataset to examine the quality of care for pregnant women with acute malnutrition and anemia in Ethiopia.

**Methods and findings:** We analyzed data from the Maternal and Newborn Health eCohort, a multi-country longitudinal study of the content, quality, and experience of maternity care during the prenatal, delivery, and postnatal periods. Data were collected in East Shewa in the Oromia region of Ethiopia from April 2023 to February 2024. Participants were recruited by research staff in a sample of health facilities at the time of their first antenatal care (ANC) contact and followed through delivery. Following Ethiopia’s national guidelines, acute malnutrition was defined as mid-upper arm circumference (MUAC) under 23 centimeters (cm), and anemia was defined as Haemoglobin (Hb) under 11 grams per deciliter (g/dL). We measured adherence to Ethiopia’s national guidelines for screening, prevention, and management of acute malnutrition, and conducted a longitudinal analysis using generalized estimating equations (GEE) to assess factors associated with adherence to iron and folic acid supplementation.

1000 women were enrolled in the eCohort. After excluding women who were lost to follow-up (n= 113), had miscarriages (n=29), or had missing information on nutritional status at baseline (n = 48), the analytic sample included 810 women. At the time of their first ANC contact, 22% of women in the sample had acute malnutrition and 11% had anemia. During the first ANC contact, only 12% of women were screened for acute malnutrition and 96% were screened for anemia. In follow-up care, weight monitoring according to national guidelines was performed for only 4% of women with acute malnutrition, and only 23% of women with anemia received the recommended number of blood tests. Only 11% of women with acute malnutrition ever received supplemental food during their pregnancy. While 89% of women with anemia were given or prescribed iron and folic acid supplements at some point during their pregnancy, only 32% took them throughout the entire pregnancy. Women with secondary education, women in the highest wealth quintile, women who reported past pregnancy complications, and women who received or were prescribed IFA supplements during the first ANC contact were more likely to adhere to IFA supplementation during pregnancy.

**Conclusions:** This study sheds light on striking gaps in the quality of care for undernutrition during pregnancy in Ethiopia. It is critical to identify approaches to improve the quality of care for undernutrition during pregnancy, focusing not only on screening and management at the first ANC contact but also on follow-up care through the remainder of pregnancy.

## INTRODUCTION

Undernutrition during pregnancy, including both micronutrient deficiencies and chronic energy deficiency, is an important public health concern with negative consequences for maternal, neonatal, and child health, development, and economic outcomes through the life course (1,2). Suboptimal nutrition during pregnancy places women at a higher risk of infections due to a compromised immune system and at a higher risk for obstetric complications, such as postpartum hemorrhage and pre-eclampsia (3–5). For the developing fetus, maternal undernutrition is linked to intrauterine growth restriction, leading to higher risk of stillbirth, low birth weight, and preterm birth (6,7). The effects of maternal undernutrition extend beyond infancy to cognitive development, educational attainment, income, and susceptibility to infectious and chronic diseases later in life, thus perpetuating a cycle of poor health and socioeconomic disadvantage (8–11).

Worldwide, an estimated 10% of women of reproductive age are underweight and 30% have anemia (12). Women are considered underweight if they have a Body Mass Index (BMI) under 18.5 kilograms per meter squared. However, during pregnancy, BMI varies with gestational age and many women start antenatal care after their first trimester; as a result, in some settings (including in Ethiopia) mid-upper arm circumference (MUAC) is used instead of BMI to identify pregnant women who are undernourished (13–15). Anemia during pregnancy is defined as Hemoglobin (Hb) under 11 grams per deciliter (g/dL) of blood (14), with higher thresholds used for women living at higher elevation and lower thresholds used in the second trimester of pregnancy (16). Guidelines for management of undernutrition in pregnancy include screening during the first antenatal care (ANC) contact, weight and Hb monitoring over the course of subsequent contacts, nutritional counseling, provision of prophylactic interventions, such as iron and folic acid (IFA) supplementation, and, where needed, therapeutic interventions such as increased iron supplementation for women with anemia and supplemental foods for underweight women (6,14). Despite widespread efforts to increase coverage of these interventions, there is little evidence on the quality of management of maternal undernutrition in high-burden settings.

Ethiopia has one the highest burdens of maternal undernutrition worldwide, which has been attributed to a variety of factors including insufficient dietary intake and largely plant-based diets (12,17). In 2016, an estimated 22% of women of reproductive age were underweight and 24% had anemia (18). In recent years, the malnutrition situation in Ethiopia been further exacerbated by conflict, drought, and a global food shortage (12). According to national guidelines, these women should be enrolled in a Targeted Supplementary Feeding Program (TSFP), monitored, and given supplementary foods to regain weight until their MUAC is in the normal range (>=23 cm) for two consecutive contacts (15). To prevent anemia, national guidelines recommend that all women take 60 mg elemental iron combined with 0.4 mg folic acid daily for six months (180 tabs) (19). To treat women with Hb < 11gm/dL, national guidelines recommend a higher dose of iron, more frequent testing and monitoring, and, in severe cases (Hb < 7gm/dL), referral to a hospital for additional care including a possible blood transfusion (19). Past studies have identified significant gaps in the quality of antenatal care in health facilities in Ethiopia (20–23), but research on the quality of management of acute malnutrition and anemia during antenatal care is scarce.

In this study, we examined the quality of care for pregnant women with acute malnutrition and anemia in Ethiopia. We used a novel longitudinal dataset that includes anthropomorphic and Hb measures at baseline, and longitudinal measures of care-seeking, health care quality, and treatment adherence. We aimed to (1) assess adherence to national clinical guidelines for the management of anemia and acute malnutrition throughout pregnancy, and (2) assess factors associated with adherence to IFA supplementation, a critical intervention for anemia prevention and management.

## METHODS

### Study design

We analyzed data from the Maternal and Neonatal Health eCohort (MNH eCohort), a multi-country longitudinal study of the content, quality, and experience of maternity care during the prenatal, delivery, and postnatal periods (24) for the case of Ethiopia.

### Setting

This study was conducted in East Shewa in the Oromia region of Ethiopia. The population of East Shewa includes approximately 500,000 people living in Adama Town, the administrative capital, and approximately 2.1 million people living in predominantly rural areas outside of Adama Town. East Shewa is divided into (1) Adama Town special zone and (2) East Shewa zone, comprised of the areas outside of Adama Town.

The Ethiopian health care system includes three levels of care. At the tertiary level, specialized hospitals serve populations of approximately 3.5-5.0 million. At the secondary level, general hospitals serve approximately 1-1.5 million people. At the primary level in urban areas, health centers serve approximately 40,000 people. The primary level in rural areas is comprised of health posts (service 3000-5000 people), health centers (serving 15,000-25,000 people), and primary hospitals (serving 60,000 to 100,000 people). Maternal health care services are available at all three levels of the health system; more complicated cases are referred from the primary to secondary or tertiary levels for specialized care (25).

### Participants

Participants were recruited from 10 health facilities in Adama Town and 11 health facilities in six districts in East Shewa Zone from 3 April 2023 to 29 May 2023. The sample included public primary, public secondary, and private health facilities and was done separately in the two zones. For sampling purposes, strata were defined by facility ownership and facility type (public primary, public secondary, private primary, and private secondary). Target sample sizes were defined for each stratum to align with care-seeking patterns for ANC in the 2019 mini DHS survey. Within strata, facilities were purposively selected based on service delivery volumes from the DHIS 2 over the period of July 2021 to June 2022 with the goal of meeting stratum-level sample size targets within the 2-month recruitment period. However, during data collection, volume of first ANC contacts in the private secondary facilities was too low and that strata was excluded.

Women were recruited by research staff at the health facility at the time of their first ANC contact, irrespective of gestational age. Women were eligible if they were pregnant, aged 15 years or older, attending the first ANC contact of their current pregnancy, intending to continue to receive maternal health care in the same zone, willing to be contacted on the phone every month for the duration of the study, and able to provide informed consent. For this study, we restricted our analytic sample to participants who completed post-delivery follow-up interviews (Module 3 of the MNH eCohort, completed 2-4 weeks postpartum) and had no missing information for MUAC or Hb level at baseline. We also excluded women who had miscarriages (defined as pregnancy loss before 28 weeks) because follow-up information on the care that these women received was incomplete: after sharing their pregnancy outcome, they were not asked any further questions about care received between the previous interview and the end of their pregnancy.

### Data sources

We analyzed data from health facilities assessments, a baseline in-person survey at the time of the first ANC contacts, and monthly follow-up phone surveys during pregnancy and until 2-4 weeks postpartum. The baseline survey also included physical health assessments (MUAC, weight, height and Hb test) and a review of the Integrated Maternal and Child Health Care Card, a facility-based health record used in Ethiopia. Recruitment took place from April-May 2023, and post-delivery phone interviews (Module 3 of the MNH eCohort) took place from June 2023 to February 2024.

### Measurement

Following Ethiopia’s national guidelines (19), acute malnutrition was defined as MUAC <23 cm. MUAC was measured by the study team for all women at enrollment following the first ANC contact. As a secondary measure, we also reported the proportion of women who were underweight following the definition of body-mass-index (BMI) <18.5 kilograms per meter squared, among those who started ANC in the first trimester.

Our primary measure of anemia was defined as Hb < 11g/dL, measured at the time of enrollment. If an Hb level from the first ANC contact was recorded in the woman’s Integrated Maternal & Child Health Care Card, this value was used. If not, Hb was assessed by the study team using Hemocue devices Model Hb 301. Hb from 10.0-10.9 g/dL was defined as mild anemia; Hb from 7.0-9.9 g/dL was defined as moderate anemia; and Hb <7g/dL was defined as severe anemia, per national guidelines (19). As a secondary measure, we also reported anemia prevalence adjusted for the pregnancy trimester and residential elevation, in line with recent World Health Organization (WHO) guidance (16). To account for elevation, we subtracted 1.1 g/dL from all measured Hb levels based on the average elevation of East Shewa (approximately 1,700m above sea level). Using this elevation-adjusted Hb level, we then applied the WHO’s recommended anemia thresholds based on pregnancy trimester (mild anemia as Hb 10.0-10.9 g/dL in the first and third trimester, and 9.5-10.4 g/dL in the second trimester; moderate anemia as Hb 7.0-9.9 g/dL in the first and third trimester, and 7.0-9.4 g/dL in the second trimester; and severe anemia as Hb <7g/dL in all trimesters). We did not use elevation and trimester adjustments for our primary measure because our goal was to evaluate adherence to Ethiopia’s national guidelines, which use the 11g/dL anemia threshold regardless of trimester or residential elevation (19).

We measured gestational age based on the date of women’s last menstrual period as reported at the time of their first ANC contact. If the date of the last menstrual period was unavailable, we relied on maternal estimates of gestational age as reported at the time of the first ANC contact.

We calculated household wealth quintiles using a principal component analysis of household asset ownership variables (ownership of a radio, television, telephone, refrigerator, car, motorbike, bicycle; having a bank account; having access to a safe water source; having an improved toilet) and house construction (floor material, wall material, and roofing material).

### Analysis

We first described sample characteristics, including the prevalence of anemia and acute malnutrition, using data collected at the time of the first ANC contact. We also described the characteristics of the health facilities where women were enrolled in the eCohort.

We then assessed adherence to Ethiopian national guidelines for the treatment of acute malnutrition and anemia during pregnancy as summarized in **Table S1**. We measured guideline adherence based on maternal recall during the baseline interview and follow-up interviews, and – for outcomes related to record-keeping – based on the Integrated Maternal and Child Health Care Card. We first reported the quality of nutrition-related care during the first ANC contact, a critical moment for initial screening and management of acute malnutrition and anemia. We then reported the quality of nutrition-related follow-up care for the subset of women with acute malnutrition or anemia at the time of the first ANC contact.

We grouped indicators into six categories: (1) screening, (2) prevention, (3) counseling, (4) treatment, (5) monitoring, and (6) record-keeping (**Table S2**). We define anemia screening based on maternal self-report of any blood test (either by a finger prick or blood draw) at the time of the first ANC contact, since both rapid test and venous blood draws are used for anemia testing in health facilities in Ethiopia. This measure may be biased upwards as some women who report blood tests may not have had Hb tests. To check this, we also review whether Hb levels were recorded in women’s Maternal and Child Health Care Cards at the time of their first ANC contact. Monitoring indicators were focused on weight monitoring for women with acute malnutrition at baseline, and Hb monitoring for women with anemia at baseline. Our analysis of adherence to monitoring guidelines was focused on the number of checks that are recommended for all women in Ethiopia’s national guidelines: 8 weight checks (at the first ANC contact and at 20, 26, 30, 34, 36, 38, and 40 weeks of gestation) and 3 blood tests (at the first ANC contact and at 30 and 38 weeks of gestation). This should be considered a minimum, as the guidelines recommend that women with acute malnutrition or anemia receive more frequent monitoring than other women, based on the progression of their condition (15,19). As a secondary analysis, we also plotted the average cumulative number of weight measures (for women with acute malnutrition at baseline) and blood tests (for women with anemia at baseline) by gestational age. Our analysis of weight and Hb monitoring is likely to be biased upwards due to a feature of our data collection: during follow-up interviews throughout pregnancy, women were asked how many routine ANC contacts they had had since their last follow-up interview, whether they had had at least one weight measurement since their last follow-up interview, and whether they had had at least one blood test (finger prick or blood draw) since their last follow-up interview. If women reported that they had had at least one weight measurement (or blood test) since their last follow-up interview, we assumed that they had had their weight measured (or blood tested) at every routine antenatal care consultation since their last follow-up interview. Since women are enrolled at the time of their first ANC contact, we assume no testing was done prior to enrolment.

Our analysis of prevention and treatment interventions was focused on the main nutritional interventions for acute malnutrition and anemia (supplemental food and IFA supplementation, respectively). In secondary analyses, we also reported the coverage of other nutritional interventions (multivitamin supplements, calcium supplements, vitamin A supplements) and of other interventions that can potentially affect anemia (malaria chemoprophylaxis, insecticide-treated bed nets, deworming medication).

Finally, we analyzed adherence to IFA supplementation using data from follow-up phone interviews throughout pregnancy. We plotted mean IFA adherence by gestational age from 20 to 40 weeks. As a secondary analysis, we plotted the same figure, restricting the sample to women who started ANC at 20 weeks or earlier, to follow the same women over time in the figure. We then conducted a longitudinal analysis of the associations between maternal characteristics and adherence to IFA supplementation using data from monthly phone interviews after ANC1. We used generalized estimating equations (GEE) to account for within-subject correlation over time (26). Our outcome was a binary variable for IFA adherence, defined as responding “yes” to the question, “Are you currently taking iron and folic acid pills?” and “everyday” to the question, “How often do you take iron and folic acid pills?”

Our main model used a logit link function and an exchangeable correlation structure. It included fixed effects for the health facility used for the first ANC contact to adjust for facility-and community-level confounders of the association between maternal characteristics and IFA adherence. It also included the following woman-level time-invariant characteristics: time since first ANC, gestational age at first ANC, anemia at first ANC (none, mild, moderate, or severe), age at first ANC, formal employment, education level, first pregnancy, past pregnancy complications (an indicator for past miscarriage, stillbirth, or neonatal death), comorbidities (an indicator for having diabetes, high blood pressure, cardiovascular disease, kidney disease, HIV, or a mental health disorder prior to pregnancy), wealth quintile, and a variable for whether she was prescribed or given IFA supplements at first ANC. We distinguish between being prescribed supplements and being given them directly because, when the supplements given in the facility are typically free while prescribed supplements are typically not free.

We conducted several sensitivity analyses. To assess associations between facility characteristics and IFA adherence, we conducted an analysis incorporating facility characteristics instead of facility fixed effects. To account for potential bias from differential follow-up periods (due to varying enrollment times based on when women begin ANC during their pregnancy), we conducted a sensitivity analysis with a restricted sample. The restricted sample included women who began ANC at 20 weeks or earlier, and only used data from follow-up interviews from 20 weeks or later, in order to follow a consistent cohort. To assess bias in quality of care measurement by omitting women who went on to experience miscarriage, we compared quality of care at first ANC among women who experienced a miscarriage with women who did not. Finally, to assess the sensitivity of our results to our choice of model specification, we conducted an analysis using a linear GEE.

### Ethical review

The protocol for this study was reviewed and approved by the Institutional Review Boards of the Harvard T. H. Chan School of Public Health (protocol #IRB22-0487) and the Ethiopia Public Health Institute (protocol number EPHI-IRB-448-2022).

## RESULTS

1000 women were enrolled in the Ethiopia eCohort. After excluding women who were lost to follow-up (n= 113), had miscarriages (n=29), or had missing information on nutritional status at baseline (n = 48), the analytic sample included 810 women (**Figure S1**). In this sample, 174 (21.5%) of women had acute malnutrition (MUAC <23cm) and 90 (11.1%) had anemia (Hb < 11 g/dL) at the time of their first ANC contact (**Table 1**). Among those with anemia, 73 (81%) had mild anemia (Hb < 11 g/dL and >=10 g/dL) and 17 (19%) had moderate anemia (Hb <10g/dL and >= 7 g/dL). No participants had severe anemia (Hb < 7 g/dL). East Shewa Zone had a higher prevalence of both anemia (18.1% vs. 3.8%) and underweight (24.9% vs. 17.9%) than Adama Town. Among the 235 women who started ANC in the first trimester, 54 (23.0%) had BMI<18.5. When using the elevation- and trimester-adjusted measure to identify women with anemia, the estimated anemia prevalence was 23.1% (14.2% mild, 8.7% moderate, and 0.1% severe).

**Table 1:** Characteristics of study participants by nutritional status.

| Characteristic | All<br>(N=810) | Acute malnutrition<br>(MUAC<23cm)<br>(N=174) | Anemia<br>(Hb <11g/dL)<br>(N=90) |
| --- | --- | --- | --- |
|  | % or Mean (SD)<sup>4</sup> | % or Mean (SD) | % or Mean (SD) |
| **Location** |  |  |  |
| Study area |  |  |  |
| Adama | 48.9% | 40.8% | 16.7% |
| East Shewa | 51.1% | 59.2% | 83.3% |
| Facility type for first ANC contact |  |  |  |
| Public Primary | 63.6% | 67.8% | 54.4% |
| Public Secondary | 7.2% | 7.5% | 15.6% |
| Private Primary | 29.3% | 24.7% | 30.0% |
| **Demographic and health characteristics** |  |  |  |
| Age | 25.5 (4.8) | 24.0 (4.4) | 24.7 (5.1) |
| Number of pregnancies (including current) | 2.5 (1.4) | 2.1 (1.1) | 2.9 (1.9) |
| Height (centimeters) | 159.5 (5.9) | 158.5 (6.1) | 158.9 (5.4) |
| Weight (kilograms) | 55.9 (10.1) | 47.8 (6.2) | 52.9 (7.5) |
| Prior obstetric complication | 8.4% | 5.7% | 7.8% |
| Chronic health condition<sup>2</sup> | 8.0% | 9.2% | 4.4% |
| **Socioeconomic characteristics** |  |  |  |
| Formal employment<sup>3</sup> | 19.4% | 17.8% | 14.3% |
| Highest level of education completed |  |  |  |
| Less than primary | 38.1% | 39.7% | 41.1% |
| Primary | 40.0% | 38.5% | 40.0% |
| Secondary or higher | 21.9% | 21.8% | 18.9% |
| Health insurance | 19.0% | 18.4% | 25.6% |
| Wealth quintile<sup>4</sup> |  |  |  |
| 1 (Poorest) | 20.1% | 27.3% | 29.2% |
| 2 | 20.1% | 22.7% | 19.1% |
| 3 | 21.3% | 26.7% | 22.5% |
| 4 | 18.8% | 14.0% | 11.2% |
| 5 (Richest) | 19.8% | 9.3% | 18.0% |
| **Antenatal care** |  |  |  |
| Trimester at 1<sup>st</sup> ANC contact |  |  |  |
| First trimester | 29.6% | 33.7% | 18.0% |
| Second trimester | 63.7% | 60.4% | 68.5% |
| Third trimester | 6.7% | 5.9% | 13.5% |
| Received at least 4 ANC contacts | 74.4% | 76.0% | 73.3% |

**Notes:** <sup>1</sup>"Prior obstetric complications" is defined as 1 if the participant has had a miscarriage, premature birth, or neonatal mortality in the past, and 0 otherwise. This variable is coded as 0 for participants with no past pregnancies. <sup>2</sup>"Chronic health condition" is defined as 1 if the participant reported that, prior to her pregnancy, she was diagnosed with diabetes, high blood pressure, cardiovascular disease, a mental health disorder, HIV, Hepatitis B, or a kidney disorder. <sup>3</sup>Formal employment includes employment with a government agency, non-governmental organization, or private company. <sup>4</sup>Wealth quintiles are calculated based on a principal component analysis information on household assets. <sup>5</sup>Standard deviation.

At the time of enrollment, 29.6% of women were in their first trimester, 63.7% were in their second trimester, and 6.7% were in their third trimester. The average age in the sample was 25.5 years (standard deviation (SD) 4.8), and the average number of pregnancies (including the current pregnancy) was 2.5 (SD 1.4). 18.4% of women with acute malnutrition and 25.6% of women with anemia had some form of health insurance. Women who had anemia at baseline were more likely to attend a public secondary facility (15.6%) and less likely to attend a public primary facility (54.4%). Of the 18 facilities in the sample, 6 offered Hb testing on-site and 10 offered Hb testing off-site (**Table S3**).

During the first ANC contact, the proportion of women having their weight measured and getting a blood test was high (81.1% and 96.0%, respectively), but only 11.7% were screened for acute malnutrition using MUAC and only 2.7% had their height measured (**Table 2**). Most women (80.9%) were given or prescribed IFA supplements. Among women with a MUAC < 23cm as measured by the study team at enrollment (acutely malnourished) only 15.5% reported that their provider had measured their MUAC during their ANC contact, and only 6.4% were given supplemental food. About one third of women (34.5%) received nutritional counseling during their first ANC contact: this was slightly higher for those with acute malnutrition (39.1%) but not for those with anemia (34.4%). Among women with anemia, only 30.0% were aware of their anemia status during their interviews immediately following the contact. Quality of care at ANC1 was similar in the analytic sample and among women who went on to experience miscarriage (who were eventually omitted from the analytic sample). The only statistically significant difference between these groups was in the portion of women who had any blood testing at first ANC contact (96.0% in the analytic sample compared with 87.0% in the miscarriage sample; p-value .03) (**Table S4**).

**Table 2:**
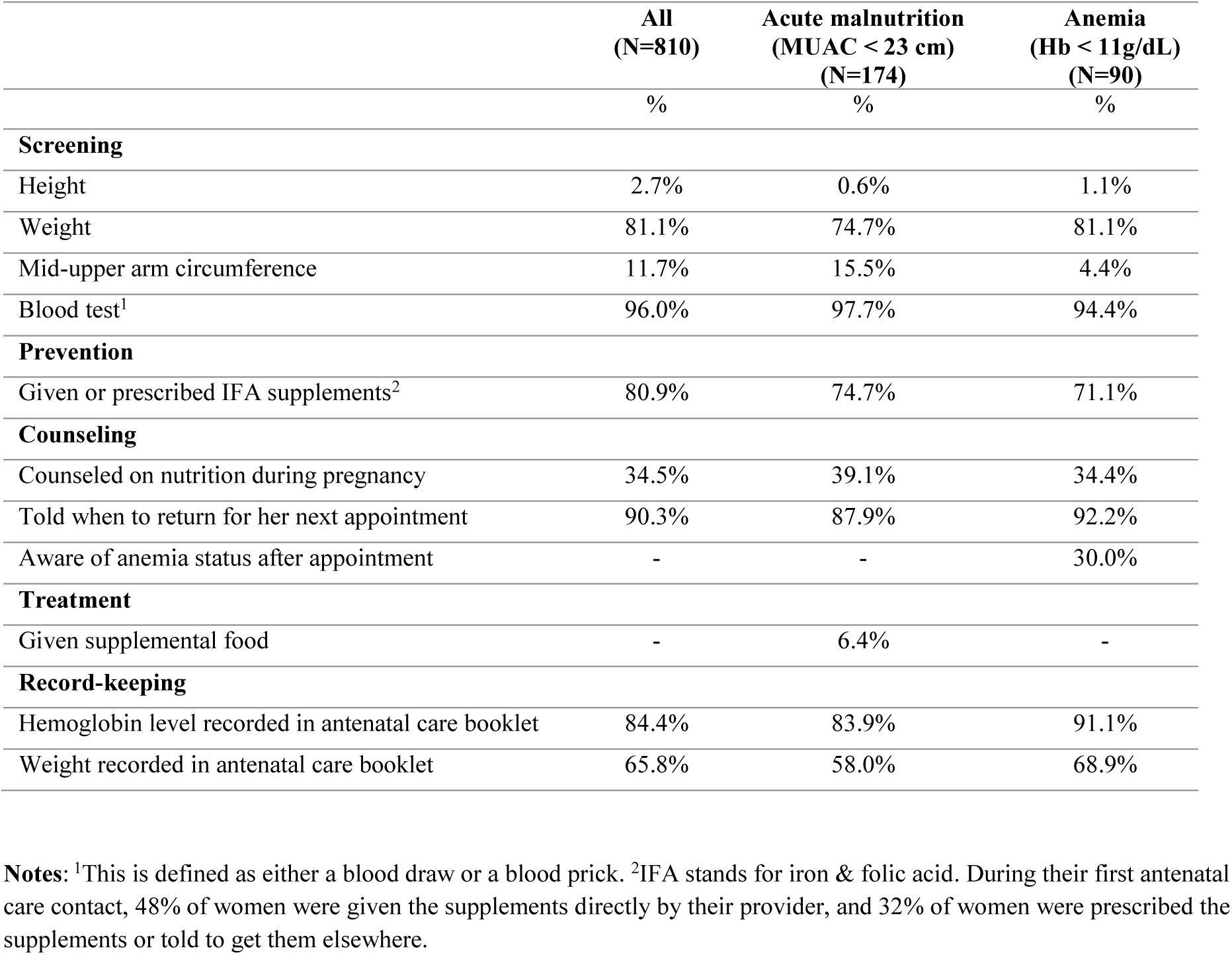
Screening, prophylactic interventions, and therapeutic interventions for anemia and acute malnutrition during the first antenatal care contact.

|  | All<br>(N=810) | Acute malnutrition<br>(MUAC < 23 cm)<br>(N=174) | Anemia<br>(Hb < 11g/dL)<br>(N=90) |
| --- | --- | --- | --- |
|  | % | % | % |
| **Screening** |  |  |  |
| Height | 2.7% | 0.6% | 1.1% |
| Weight | 81.1% | 74.7% | 81.1% |
| Mid-upper arm circumference | 11.7% | 15.5% | 4.4% |
| Blood test<sup>1</sup> | 96.0% | 97.7% | 94.4% |
| **Prevention** |  |  |  |
| Given or prescribed IFA supplements<sup>2</sup> | 80.9% | 74.7% | 71.1% |
| **Counseling** |  |  |  |
| Counseled on nutrition during pregnancy | 34.5% | 39.1% | 34.4% |
| Told when to return for her next appointment | 90.3% | 87.9% | 92.2% |
| Aware of anemia status after appointment | - | - | 30.0% |
| **Treatment** |  |  |  |
| Given supplemental food | - | 6.4% | - |
| **Record-keeping** |  |  |  |
| Hemoglobin level recorded in antenatal care booklet | 84.4% | 83.9% | 91.1% |
| Weight recorded in antenatal care booklet | 65.8% | 58.0% | 68.9% |

**Notes:** <sup>1</sup>This is defined as either a blood draw or a blood prick. <sup>2</sup>IFA stands for iron & folic acid. During their first antenatal care contact, 48% of women were given the supplements directly by their provider, and 32% of women were prescribed the supplements or told to get them elsewhere.

During follow-up ANC contacts, women with acute malnutrition and anemia received very little monitoring (**Figure 1**). Only 4% of women with acute malnutrition at baseline received the recommended number of weight checks during pregnancy, and only 23% of women with anemia at baseline received the recommended number of blood tests. Among women with acute malnutrition at baseline, we estimate an average cumulative number of weight checks of 1.4 at 20 weeks gestation, and 3.3 at 40 weeks gestation (**Figure S4, Panel A**). This increase of less than 2 weight checks over 20 weeks of gestation indicates that women with acute malnutrition receive less than the recommended frequency of weight checks for women at a healthy weight.

**Figure 1:**
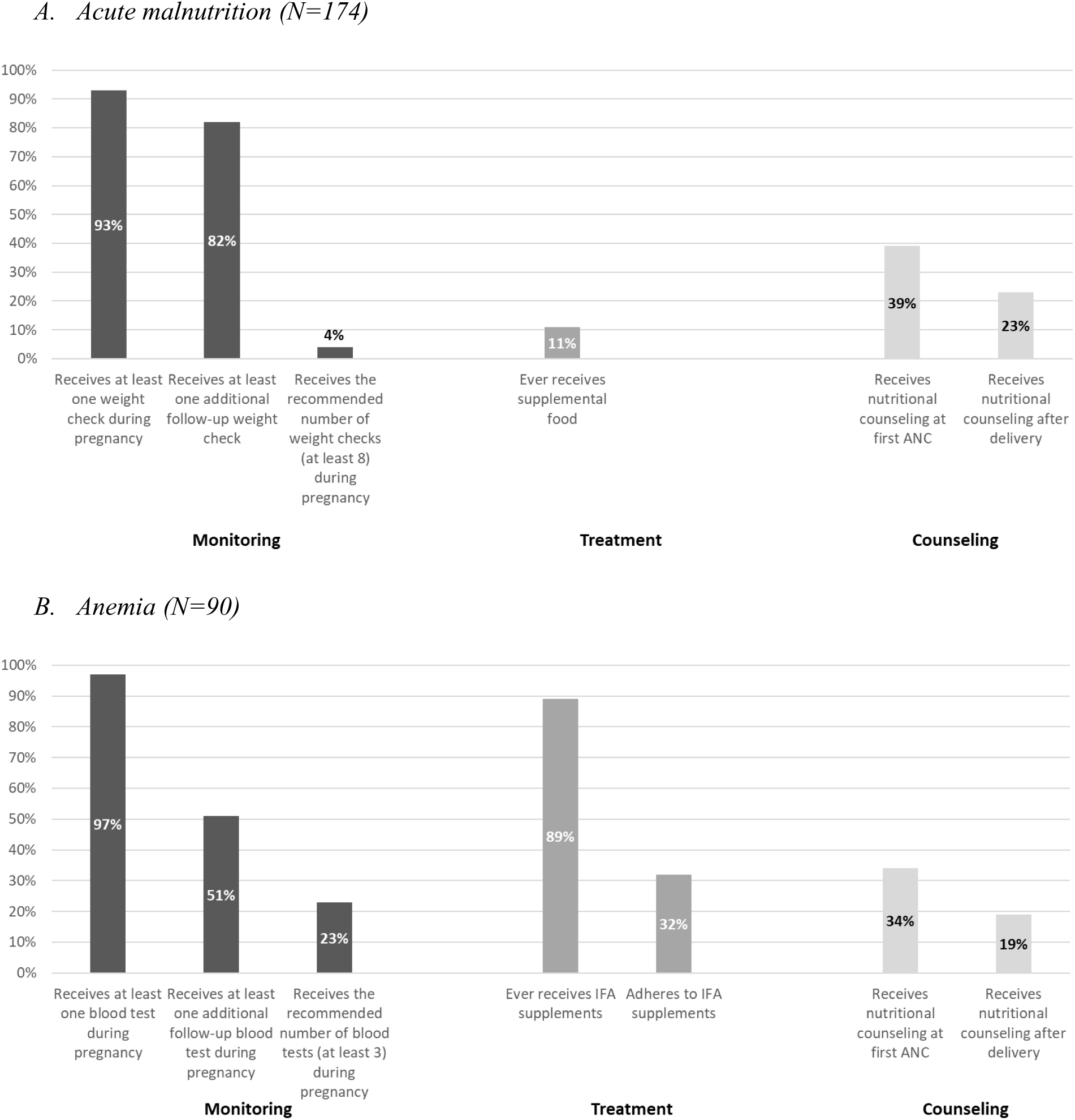
Follow-up antenatal care for women with acute malnutrition and anemia. **Notes**: ANC stands for antenatal care. Hb stands for Haemoglobin. Dark gray bars indicate care received during the first antenatal care contact, as reported by participants during baseline face-to-face interviews. Medium gray bars indicate care received during follow-up antenatal care contacts, as reported by participants during monthly phone interviews. Light gray bars indicate care received at the time of delivery, as reported during phone interviews taking place approximately two weeks after delivery.

**Figure 2:**
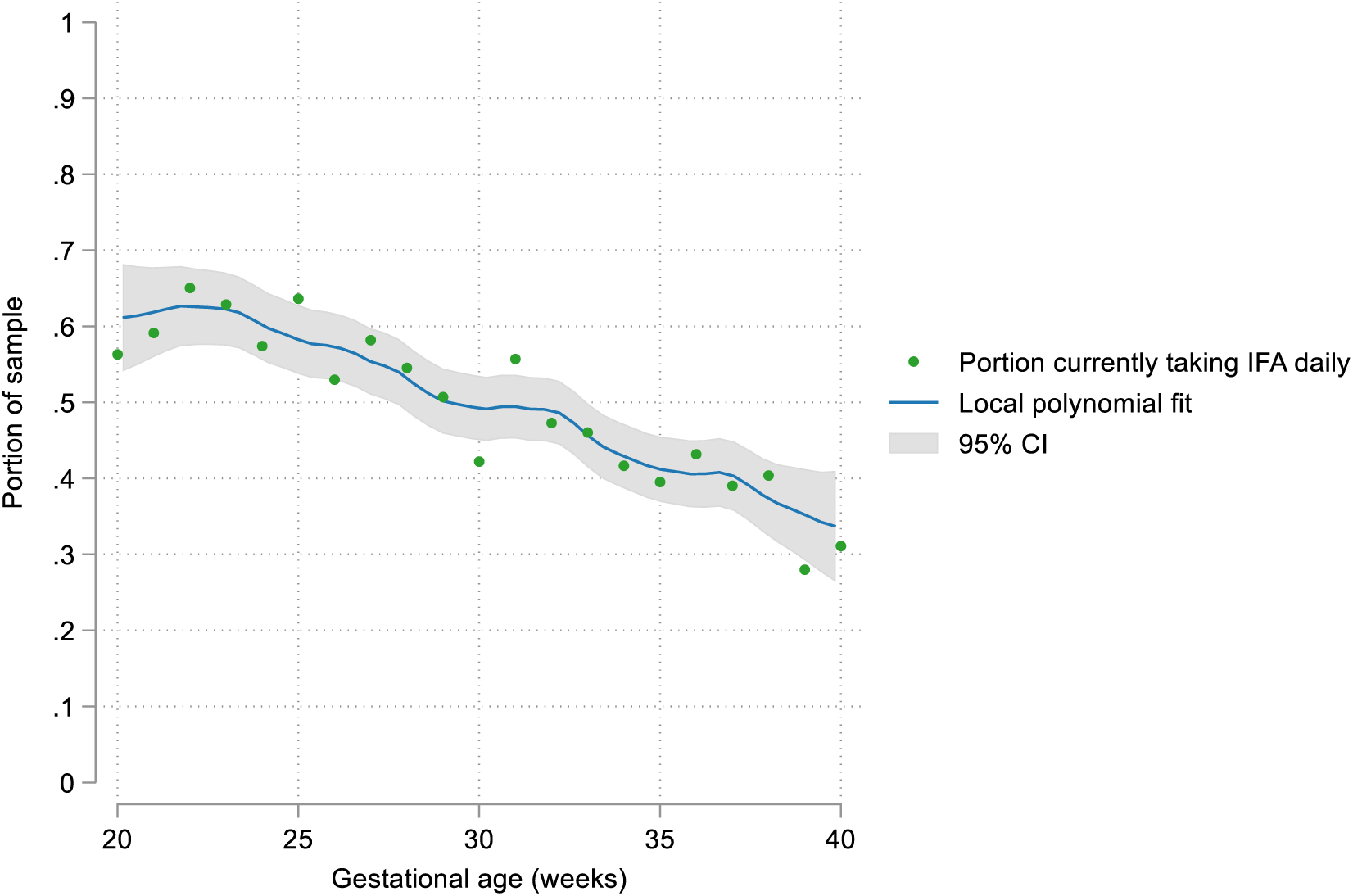
Self-reported daily adherence to iron and folic acid supplementation during pregnancy, by gestational age. **Notes**: This figure shows data from monthly follow-up phone interviews throughout pregnancy. Gestational age at the time of the interview is rounded to the nearest week. Green dots show the proportion of women who, during interviews at each gestational age, reported daily adherence to iron & folic acid supplementation. The blue line shows an estimated local polynomial fit estimated using a kernel density with an Epanechnikov kernel, and the gray shading shows the 95% confidence interval around the line.

Treatment coverage was also low. Only 11% of women with acute malnutrition ever received supplemental food during the pregnancy (including during the first ANC contact and follow-up contacts). While 89% of women with anemia received IFA supplements at some point during pregnancy, only 32% of them reported daily adherence throughout pregnancy. Post-delivery nutritional counseling was rare: only 23% of women with acute malnutrition at baseline and 19% of women with anemia at baseline received nutritional counseling after giving birth.

The coverage of other nutritional supplements is shown in **Table S5**. 17.5% of women were ever given or prescribed multivitamins. This was typically in addition to, rather than instead of, IFA supplements: 16.0% of women received both multivitamins and IFA supplements, and only 1.5% of women received multivitamins but no IFA supplements. 1.9% of women were given or prescribed calcium supplements. Among those living in a malaria-endemic area, 12.2% were given an insecticide-treated bed net during their first ANC contact, 2.4% were counseled on sleeping under an insecticide-treated bed net, and 5.4% were given or prescribed antimalarial medication. Among those living in an area with endemic helminth infections, 26.4% were given or prescribed deworming medication at some point during pregnancy.

The level of adherence to nutritional guidelines was similar for those with BMI<18.5 and those with MUAC<23 (**Table S6**). It was also generally similar for women with anemia defined by Hb < 11g/dL and for women with anemia defined by the elevation- and trimester-adjusted measure (**Table S6**). One exception was IFA adherence, which was lower when using the elevation- and trimester-adjusted anemia measure (25% vs. 32%) While nearly all (96.0%) women in the sample received IFA supplements at some time in their pregnancy, adherence to IFA supplements decreased over the course of pregnancy (**Figure 1**). The proportion of women who reported daily IFA adherence decreased from 59% at 20 weeks gestational age to approximately 33% at 40 weeks. This result was similar when the sample was restricted to women who began antenatal care before 20 weeks gestation, decreasing from 59% at 20 weeks to 27% at 40 weeks (**Figure S3**).

**Table 3** show the results from a GEE model analyzing the association between maternal characteristics and daily IFA adherence. We found that the odds of daily adherence was 25% lower for each additional 4 weeks following the first ANC contact (Odds Ratio (OR) 0.75, 95% CI: 0.71, 0.79). In the linear model, this translated to a 6 percentage point decrease (95% CI: 5, 7) for each additional 4 weeks of gestation (**Table S7**). Women with secondary education or more had higher odds of adherence (OR 1.47, 95% CI: 1.02, 2.11) when compared with women with no education. Women in the highest wealth quintile also had higher odds of adherence (OR 1.68, 95% CI: 1.09, 2.60) than women in the lowest wealth quintile. Women who reported past pregnancy complications were also more likely to adhere (OR 1.50, 95% CI: 1.08, 2.09). Receiving IFA supplements at the first ANC contact was significantly associated with adherence: the odds of adherence among women who were given supplements directly during the first ANC contact were 52% times higher (OR 1.52, 95% CI: 1.05, 2.20) compared with women who are not given or prescribed IFA supplements. The odds of adherence were substantially higher (OR 3.22, 95% CI: 1.98, 5.56) among women prescribed IFA supplements. Age, employment, chronic conditions, first pregnancy, depression at the time of the first ANC contact, and anemia level were not significantly associated with adherence. In sensitivity analysis with facility characteristics, women who attended facilities in urban areas had a 60% lower odds of adhering (OR 0.40, 95% CI: 0.28, 0.56) than who attend facilities in rural areas, and women who attended public primary facilities had a higher odds of adhering than those who attended private primary facilities (OR 1.66, 95% CI: 1.24, 2.23) (**Table S6**), adjusting for maternal demographic characteristics, socioeconomic characteristics, and pregnancy characteristics. Results from the restricted sample and the linear regression were similar to the results from the main model.

**Table 3:** Predictors of adherence to iron and folic acid supplementation.

|  | **Odds Ratio** | **95% CI** | **P-value** |
| --- | --- | --- | --- |
| Time since ANC1 (increments of 4 weeks) | 0.75*** | 0.71,0.79 | <0.001 |
| **Demographic and socioeconomic characteristics** |  |  |  |
| Age (Ref: <20) |  |  |  |
| 20-35 | 1.10 | 0.71,1.71 | 0.66 |
| 35+ | 0.96 | 0.53,1.73 | 0.89 |
| Formally employed | 1.15 | 0.85,1.56 | 0.37 |
| Education (Ref = Less than primary) |  |  |  |
| Completed primary | 1.20 | 0.92,1.55 | 0.17 |
| Completed secondary or higher | 1.47* | 1.02,2.11 | 0.04 |
| Quintile (Ref = 1 (Poorest)) |  |  |  |
| 2 | 1.10 | 0.77,1.56 | 0.61 |
| 3 | 1.03 | 0.71,1.49 | 0.88 |
| 4 | 1.14 | 0.76,1.71 | 0.52 |
| 5 | 1.68* | 1.09,2.60 | 0.02 |
| **Health history** |  |  |  |
| Past pregnancy complications | 1.50* | 1.08,2.09 | 0.02 |
| Chronic health condition | 1.03 | 0.70,1.53 | 0.88 |
| **Current pregnancy** |  |  |  |
| First pregnancy | 0.85 | 0.65,1.09 | 0.20 |
| Gestational age at ANC1 (increments of 4 weeks) | 0.95 | 0.87,1.03 | 0.20 |
| Depression at ANC1 (PHQ9 score of 5 or above) | 0.79 | 0.61,1.04 | 0.09 |
| Anemia severity at ANC1 (Ref: No anemia) |  |  |  |
| Mild anemia | 0.84 | 0.52,1.35 | 0.47 |
| Moderate anemia | 0.56 | 0.29,1.09 | 0.09 |
| Self-reported anemia status directly after ANC1 | 1.29 | 0.95,1.75 | 0.11 |
| IFA pills at ANC1 (Ref: Neither given nor prescribed) |  |  |  |
| Given by provider | 1.52* | 1.05,2.21 | 0.03 |
| Prescribed or told to get elsewhere | 3.32*** | 1.98,5.56 | <0.001 |
| Number of observations | 2766 |  |  |
| Number of women | 758 |  |  |
| Facility fixed effects | Yes |  |  |
| Clustered standard errors | Yes |  |  |

**Notes:** This table shows estimates from a Generalized Estimating Equation (GEE) logistic regression of daily iron & folic acid supplement adherence on maternal characteristics, using data from monthly follow-up interviews. The outcome was defined as a binary variable that was 1 if a woman reported that she was taking daily iron & folic acid supplements at the time of the call and 0 otherwise. Participants each had between 1 and 7 follow-up interviews. An exchangeable correlation structure was used, with standard errors clustered at the level of the participant. The model included fixed effects for the facility used for the first antenatal care contact. CI refers to the Confidence Interval. ANC1 refers to the first antenatal care contact. PHQ9 refers to the score on the Patient Health Questionnaire-9. Ref refers to the omitted reference group. \*p<0.05; \*\*p<0.01; \*\*\*p<0.001.

## DISCUSSION

In this study, we used longitudinal data to assess adherence to national guidelines for the management of acute malnutrition and anemia and investigated the factors associated with IFA adherence during pregnancy in Ethiopia. In our sample, 21.5% of women had acute malnutrition as defined by MUAC<23cm and 11.1% of women had anemia, as defined by Hb<11g/dL. The prevalence of anemia was even higher at 23.1% when accounting for elevation. We identified important gaps in the quality of their care.

First, there were important delays in ANC initiation, with the majority of women starting ANC in their second trimester. Only 34% of women with acute malnutrition and 18% of women with anemia started ANC in their first trimester. When women with acute malnutrition women start ANC late, as they did in our sample, the fetus is likely to have experienced suboptimal nutrition in-utero for a long time and therefore has an increased risk of being born low birthweight, preterm, or small-for-gestational age (27).

Second, despite the high burden of acute malnutrition, we found extremely limited screening, monitoring, and treatment of pregnant women with acute malnutrition. Only 12% of women were screened for acute malnutrition at the first ANC contact, and only 11% of women with acute malnutrition were given supplemental food at any point during pregnancy. The average number of weight checks for women suffering from acute malnutrition was only 3.

Third, we found relatively high rates of screening for anemia (96%) and of provision of prophylactic IFA supplements at the first antenatal care contact (81%), but follow-up care was limited. Only 51% of women with anemia received any follow-up blood testing. Finally, the rate of adherence to IFA supplementation was low among all women, including the subgroup of women who had anemia at their first ANC contact. Adherence varied significantly by education and wealth levels and decreased significantly over the course of pregnancy.

Past studies of antenatal care quality in Ethiopia have generally not focused on care specifically for acute malnutrition, despite the importance of this problem (21–23,28). Our study brings attention to several major gaps in care for this vulnerable group. Improving management of acute malnutrition in pregnancy will require improving the levels of screening during ANC. Less than one in eight women with acute malnutrition had their MUAC measured at their first ANC contact. Although MUAC is recommended in ANC guidelines in Ethiopia, we found that the paper-based Integrated Maternal and Child Health Card used as an ANC record in health facilities Ethiopia did not include a section to enter the MUAC measurement or to identify undernutrition as a risk factor. One step towards improving screening could be to update the Integrated Maternal and Child Health Cards so that providers are reminded to measure, record and monitor MUAC, as this is the recommended screening measure for acute malnutrition in national guidelines but is currently not documented in health records (**Figure S5**).

The low frequency of weight monitoring that we observed is also concerning: without sufficient weight monitoring during pregnancy, it is impossible to know whether gestational weight gain is sufficient to meet the needs of the fetus. In Ethiopia, care for acute malnutrition is currently geographically targeted to high-risk areas, known as hotspot districts and towns. One district in our sample (Adami Tulu) is designated as a second-priority hotspot for acute malnutrition (29). In this area, 41% of women with acute malnourishment received supplemental food at their first ANC contact and 52% received it at some point during pregnancy. The fact that coverage of supplemental food is still so low even in a designated hotspot signals a need for significant efforts to improve the quality of ANC for pregnant women with acute malnutrition. Given the high prevalence of undernutrition among pregnant women in Ethiopia, policy makers should also consider better integration of nutrition and ANC programs at the national level.

Few past studies have focused on the content of follow-up antenatal care contacts, despite the importance of these visits for monitoring and addressing complications identified earlier in pregnancy. We found that adherence to guidelines for anemia at the first ANC contact was relatively high, but women with anemia received extremely limited follow-up care. This is in line with one past study that showed low coverage of key interventions even among women who had four antenatal care contacts (21). Our study is the only one, to our knowledge, to report on the frequency of blood testing during follow-up ANC contacts among pregnant women with anemia in Ethiopia, which is critical for monitoring treatment effectiveness and informing decisions about whether to escalate care.

Our work confirms the findings of past studies in Ethiopia and other countries in sub-Saharan Africa that have documented gaps in IFA adherence during pregnancy and socioeconomic gradients in adherence (30–35). Our study makes a unique contribution to this literature through the analysis of data from monthly follow-up interviews during pregnancy, which reveal a clear downward trend in IFA adherence over the course of pregnancy. There are a number of possible explanations for this. It is possible that women start taking supplements early in pregnancy find it too difficult to remember, or too uncomfortable due to side effects, and eventually stop taking them. Indeed, a qualitative study in Ethiopia cited women’s negative experiences with IFA supplements, and a lack of information on their benefits and risks, as key reasons for low adherence (36). It is also possible that cost and operational challenges related to supply availability play a role (36). Further research is needed to better understand the reasons for non-adherence and address them such as by identifying more tolerable formulations, addressing supply and cost barriers, or improving communication to patients, drawing on past research in this area (37–39).

Our study has several limitations. First, our study excluded women who did not access any ANC, as well as women who had miscarriages or elective abortions. These women may be particularly vulnerable; understanding the care they receive over the course of their pregnancies should be a priority for future studies. As a result of these exclusions, our study likely over-estimates the true population level adherence to national guidelines in this setting. As a sensitivity analysis, we examined the quality of care at first ANC contact among women who later had miscarriages; quality in this subset was similar to quality in our analytic sample, except that the miscarriage sample was significantly less likely to have received any blood testing. Secondly, quality measurement in our study was based on maternal recall, but women may not always accurately remember or report the care they received or their adherence to medications due to problems with recall and social desirability bias. Our measure of gestational age was based on maternal recall of the last menstrual period, which is known to be a noisy measure (40). Our reliance on maternal recall also limited our ability to measure the frequency of Hb testing: some women who report blood tests (either finger pricks or blood draws) may not have had Hb tests. However, during baseline interviews, we also reviewed Maternal and Child Health Care Cards and found that nearly all women who reported blood tests also had an Hb level in their records. Our measures of follow-up blood testing and weight monitoring also provide an upper bound on the true level of monitoring because of the way these questions were formulated in follow-up interviews. Thirdly, our sample was selected to be representative of antenatal care-seeking patterns in the two sampled health zones, but was not regionally or nationally representative. The prevalence of acute malnutrition and anemia in our sample were somewhat lower than those reported in past studies at the national and regional levels. Two recent systematic reviews of malnutrition in Ethiopia reported pooled estimates of 32% (95% CI: 31, 33) and 29% (95% CI: 25, 33) among pregnant women nationwide (41,42), and 30% (95: CI: 21, 39) among pregnant women in Oromia, with estimates in Oromia ranging from 12% to 47% using several different definitions (BMI < 18.5, MUAC < 21, MUAC < 22, MUAC < 23) (41). The 2016 DHS in Ethiopia estimated an anemia prevalence of 27% among women aged 15-49 in Oromia (using Hb cutoffs of 12g/dL among non-pregnant women and 11g/dL among pregnant women) (18), and a recent systematic review estimated a pooled prevalence of 26% (95% CI: 26, 27) among pregnant women in Oromia (using an Hb cutoff of 11g/dL) (43), both without elevation adjustment. A recent study estimated elevation-adjusted anemia prevalence among women aged 15-49 in Oromia of 16.7%, varying from 11.8% in the dry season (October to May) to 21.8% in the wet season (June to September) (17). Differences are likely driven by geographic and socioeconomic heterogeneity in samples (e.g., our sample is more concentrated in urban areas than the overall national population), seasonality (our sample was recruited during the dry season), variation in definitions of malnutrition and anemia, and the exclusion of women who had no ANC and women who experienced miscarriages from our study sample. Fourthly, our data did not include information on IFA supplement dosing, and we were thus unable to assess whether women with anemia received higher doses of IFA supplements. We also did not systematically measure the coverage of iron injections or blood transfusions, and are thus unable to report on these treatments that are required in more severe anemia cases. Finally, as the MNH eCohort was designed to study the quality of maternal health care in a general sample of pregnant women and not focused exclusively on pregnant women with nutritional deficits, the number of participants in the subsets with acute malnutrition (N=174) and anemia (N=90) are relatively small.

Our results have important implications for policy. First, there is a critical need to address gaps in care for pregnant women with acute malnutrition. Our study shows that, in our study sites, there is a significant population of pregnant women with acute malnutrition who are not reached by current programs. It is critical to increase screening for and treatment of acute malnutrition during pregnancy in this setting. A second step is to ensure that supplemental food is available wherever women with acute malnutrition are seeking ANC. For women with anemia, care at the first ANC contact is generally good but there is a critical need to improve follow-up care, including Hb monitoring, during the remainder of pregnancy. Further research is also needed to test and scale up approaches for increasing IFA supplement adherence. The Ethiopian national nutrition program set a target to increase the number of women receiving IFA supplements for 90+ days during pregnancy to 50% by 2024 (44); but increasing distribution is not enough if women do not take these supplements. For both acute malnutrition and anemia, there is a need for ANC reforms and implementation research that focus not only on the first contact but on delivering high quality care over the course of pregnancy so that complications are identified early on but also adequately managed to minimize risks to the mother and fetus. This study reveals significant gaps in the management of acute malnutrition and anemia among pregnant women in Ethiopia. It is critical to identify approaches to improve the quality of care for undernutrition during pregnancy.

## Data Availability

Analytic data in for this manuscript are available on the Harvard Dataverse: Clarke-Deelder, Emma, 2025, "Replication Data for: Screening, prevention, and management of maternal acute malnutrition and anemia in Ethiopia: evidence from a longitudinal eCohort study", https://doi.org/10.7910/DVN/JGAXNG, Harvard Dataverse, V1, UNF:6:rN8mEyGG1NUNt6nBgR9idA== [fileUNF].

## Supplementary Appendix

**Table S1:** Ethiopian Guidelines for Management of Nutrition-Related Care During Pregnancy.

| Category | Ethiopian Guidelines |
| --- | --- |
| Screening for under-nourishment at first antenatal care contact | - <u>Ask about nutritional history</u> (number of food groups and frequency of meals consumed per day, craving for unusual food type, appetite, emesis) (ANC guideline, page 12)<br>- <u>Measure weight and height</u> as a baseline for monitoring during pregnancy (ANC guideline, page 13)<br>- <u>Measure mid-upper arm circumference</u> as screening for acute malnutrition (ANC guideline, page 13)<br>- <u>Examine the conjunctiva, oral mucosa, and nail beds for pallor</u> (ANC guideline, page 13)<br>- <u>Measure hemoglobin (Hb) or hematocrit (Hct)</u> (ANC guideline, page 14) |
| Monitoring of nutrition and health status at subsequent antenatal care contacts | - <u>Measure weight</u> at weeks 20, 26, 30, 34, 36, 38, and 40 (ANC guidelines, pages 15-16)<br>- <u>Measure Hb</u> at weeks 30 and 38 (ANC guidelines, pages 15-16)<br>- <u>Measure MUAC</u> at weeks 20, 26, 30, 34, 36, 38, and 40 (ANC guidelines, pages 15-16)<br>- <u>For women with MUAC<23</u>, also monitor: presence of bilateral pitting oedema; indicate any necessary follow-up actions on the Targeted Supplemental Feeding Program Treatment and Follow-up Card (Malnutrition guidelines, page 90) |
| Prophylactic interventions for nutrition and health during pregnancy (for all women) | - <u>Nutrition counseling</u> (including: counseling on diet and micronutrient supplementation; counseling on exercise; assessment of compliance and counselling on adherence of iron, folic acid, and calcium intake during each contact during pregnancy; counseling on consumption of at least one additional diversified and nutrient-dense meal on a daily basis; and promotion of consumption of adequately iodized salt intake) (ANC guidelines, page 22)<br>- <u>Iron folic acid supplementation</u> (all pregnant women should take 60mg elemental iron and 0.4mg folic acid daily for six months; 180 tabs) (ANC guidelines, page 23)<br>- <u>Calcium supplementation</u> (all pregnant women should take 1.5-2.0 gm oral elemental calcium daily from 14 weeks gestation) (ANC guidelines, page 23)<br>- <u>De-worming treatment</u> for pregnant women in their 2<sup>nd</sup> or 3<sup>rd</sup> trimester (400mg of albendazole or 500mg of mebendazole) (ANC guidelines, page 3)<br>- <u>Antimalarial medication</u> for pregnant women in their 2<sup>nd</sup> trimester (Sulfadoxine-pyrimethamine) on a monthly basis, if in a malaria endemic area (ANC guidelines, page 27) |
| Treatment interventions for undernutrition (for women who screen positive) | - For women with Hb levels <11:<br>o Mild anemia (Hb 9-10.9 gm/dl): therapeutic iron dose + peripheral RBC morphology and RBC indices<br>o Moderate anemia (Hb 7-8.9 gm/dl): Therapeutic iron dose + peripheral RBC morphology and RBC indices, close follow-up<br>o Severe anemia (Hb <7gm/dl): Referral to a hospital for complete investigation and possible blood transfusion; continue therapeutic iron dose then after |
|  | ○ If no adequate response to therapeutic iron dose, refer to a hospital for a complete investigation.<br>○ Therapeutic iron dose is: 60mg elemental iron, BID in 24 hours until the Hb rises to $\geq 11$ gm/dl, to be followed by prophylactic dose.<br>- For women with MUAC < 23:<br>○ Admit to Targeted Supplemental Feeding Program (TSFP) (Malnutrition guidelines, page 85)<br>○ Provide fortified blended food (FBF), such as Super Cereal Plus or ready-to-use supplementary food (USF) (Malnutrition guidelines, page 88)<br>○ Provide health, nutrition, and hygiene education (Malnutrition guidelines, page 89) |

Notes: Ethiopia guidelines were extracted from the 2022 National Antenatal Care Guideline and the National Guideline for the Management of Malnutrition in Ethiopia.

**Table S2:** Indicators of adherence to Ethiopia’s national guidelines for acute malnutrition and anemia management during pregnancy.

| Category | Indicator | Definition | Data source | Measurement period |
| --- | --- | --- | --- | --- |
| (1) Screening | (a) MUAC measured | Participant reports that her MUAC was measured | Maternal recall | First ANC<sup>1</sup> contact |
|  | (b) Height measured | Participant reports that her height was measured | Maternal recall | First ANC contact |
|  | (c) Weight measured | Participant reports that her weight was measured | Maternal recall | First ANC contact |
|  | (d) Blood test | Participant reports either a blood prick or blood draw | Maternal recall | First ANC contact |
| (2) Prevention | (a) Iron & folic acid supplements given or prescribed | Participant reports that she was given or prescribed IFA supplements | Maternal recall | First ANC contact |
| (3) Counseling | (a) Nutritional counseling at first ANC | Participant reports that she received any nutritional counseling during her first ANC contact | Maternal recall | First ANC contact |
|  | (b) Told when to return | Participant reports that she was told when to return for her second ANC contact | Maternal recall | First ANC contact |
|  | (c) Told anemia status | Following her first ANC contact, participant reports that she has anemia (among participants who had Haemoglobin < 11 grams per deciliter at the time of their first ANC contact) | Maternal recall | First ANC contact |
|  | (d) Postpartum nutritional counseling | Participant reports that she received counseling on nutrition following her delivery | Maternal recall | Immediately following delivery |
| (4) Treatment | (a) Ever given or prescribed iron & folic acid | Participant reports at least once during pregnancy that she was given or prescribed iron & folic acid supplements | Maternal recall | Any time during pregnancy |
|  | (b) Ever given or prescribed supplemental food | Participant reports at least once during pregnancy that she was given or prescribed supplemental food such as Plumpynut | Maternal recall | Any time during pregnancy |
| (5) Monitoring | (a) At least one weight check during pregnancy | Participants reports at least once during pregnancy that her weight was measured during a routine antenatal care contact | Maternal recall | Any time during pregnancy |
|  | (b) At least one follow-up weight check during pregnancy | Participant reports at least twice during pregnancy that her weight was measured during a routine antenatal care contact | Maternal recall | Any time during pregnancy |
|  | (c) Recommended number of weight checks during pregnancy | Participant reports at least 8 times during pregnancy that her weight was measured during a routine antenatal care contact | Maternal recall | Any time during pregnancy |
|  | (d) At least one blood test during pregnancy | Participant reports at least once during pregnancy that she received a blood test using either a finger prick or a blood draw | Maternal recall | Any time during pregnancy |
|  | (e) At least one follow-up blood test during pregnancy | Participant reports at least twice during pregnancy that she received a blood test using either a finger prick or a blood draw | Maternal recall | Any time during pregnancy |
|  | (f) Recommended number of blood tests during pregnancy | Participant reports at least three times during pregnancy that she received a blood test using either a finger prick or a blood draw | Maternal recall | Any time during pregnancy |
| (6) Record-keeping | (a) Hemoglobin level recorded in Integrated Maternal and Child Health Care Card |  | Integrated Maternal and Child Health care Card | First ANC contact |
|  | (b) Weight recorded in Integrated Maternal and Child Health Care Card |  | Integrated Maternal and Child Health care Card | First ANC contact |

<sup>1</sup>ANC = antenatal care.

**Figure S1:**
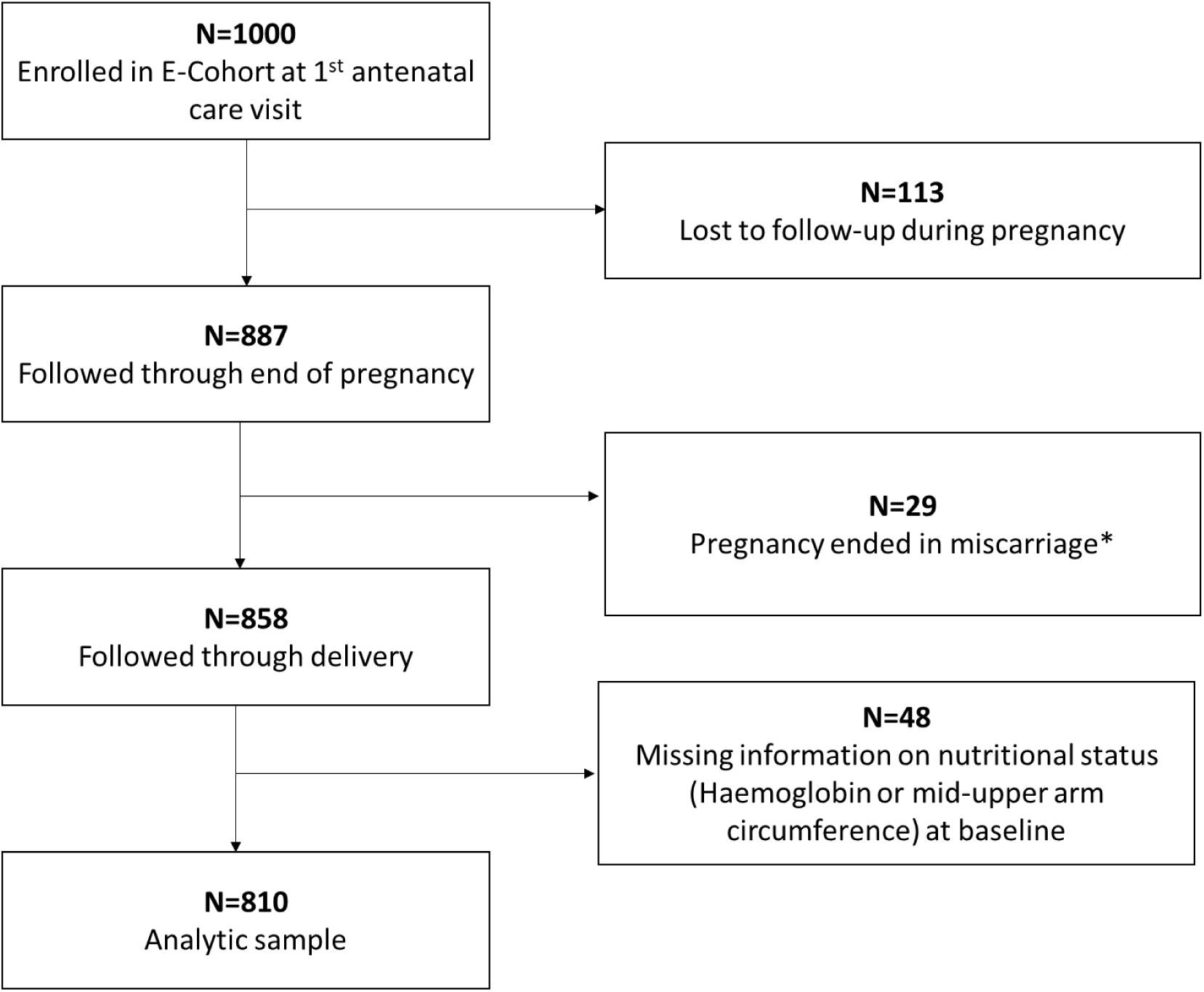
Sample flowchart. * 4 of the 29 miscarriages occurred after 28 weeks of gestation and should have been classified as stillbirths by data collectors.

**Table S3:**
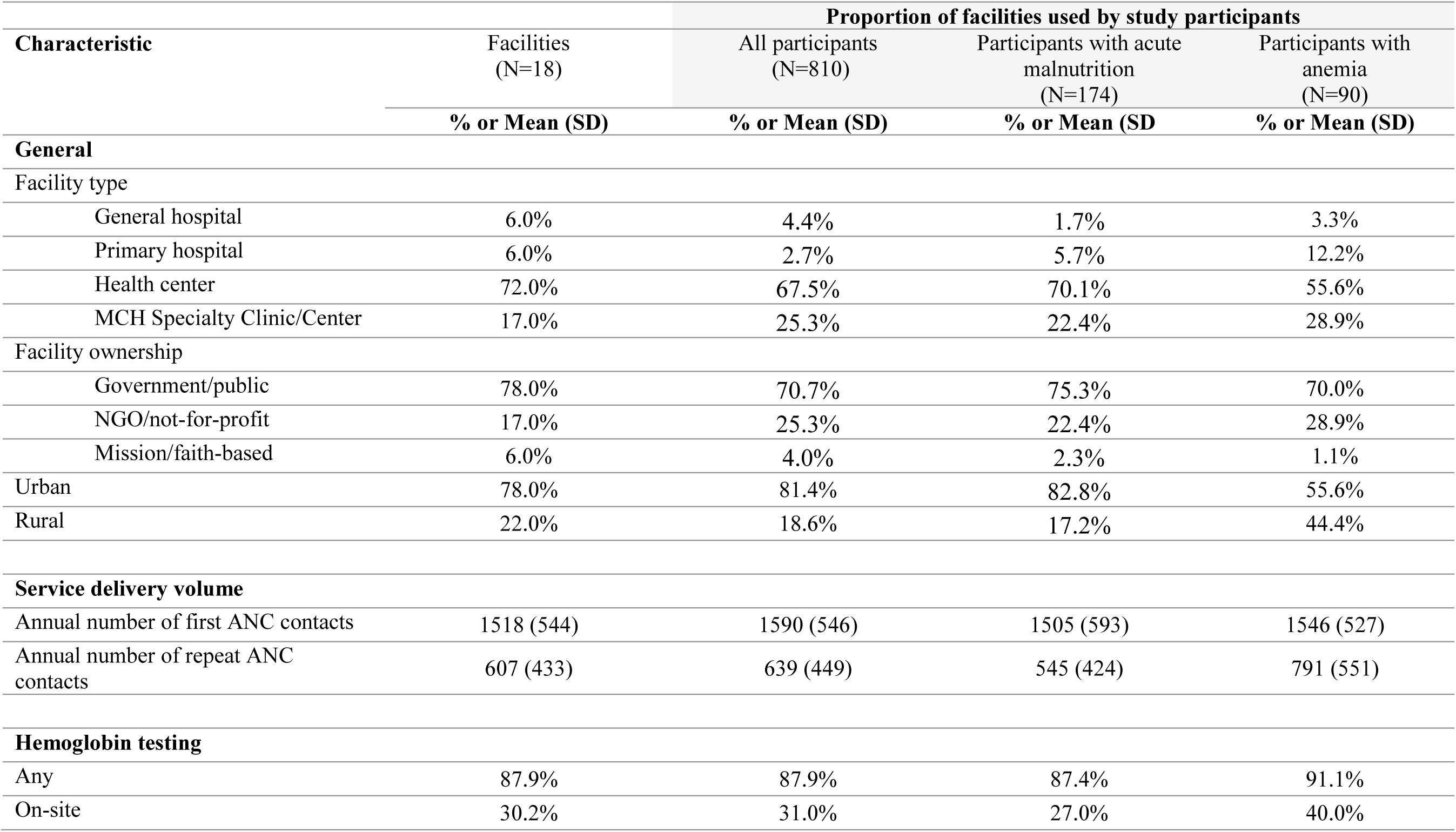
Characteristics of health facilities used for the first antenatal care contact.

**Table S4:** Additional nutrition-related interventions during pregnancy.

|  | **All**<br>**(N=810)** | **Acute**<br>**malnutrition**<br>**(N=174)** | **Anemia**<br>**(N=90)** |
| --- | --- | --- | --- |
| **First ANC<sup>1</sup> contact** |  |  |  |
| Given or prescribed multivitamins | 7.5% | 7.6% | 8.9% |
| Given or prescribed both multivitamins and iron & folic acid supplements | 4.5% | 5.8% | 4.4% |
| Given or prescribed calcium supplements (if 14+ weeks gestational age)<sup>2</sup> | 0.6% | 0.0% | 1.1% |
| Given or prescribed deworming medication, if 13+ gestational age and in an endemic area | 0.4% | 0.0% | 0.0% |
| Given or prescribed antimalarial medication, if 13+ weeks of gestational age and in an endemic area | 0.0% | 0.0% | 0.0% |
| Given an antimalarial bed net, if in a malaria endemic area | 12.2% | 11.2% | 8.4% |
| Counseled on sleeping with an antimalarial bed net, if in a malaria endemic area | 2.4% | 1.8% | 0.0% |
| **Ever during pregnancy** |  |  |  |
| Given or prescribed multivitamins | 17.5% | 19.6% | 19.1% |
| Given or prescribed both multivitamins and iron & folic acid supplements | 16.0% | 19.6% | 14.6% |
| Given or prescribed calcium supplements | 1.9% | 1.8% | 3.4% |
| Given or prescribed deworming medication, if in an endemic area | 26.4% | 26.9% | 26.1% |
| Given or prescribed antimalarial medication, if in an endemic area | 5.4% | 5.6% | 12.5% |

**Notes:**<sup>1</sup>Antenatal care

**Table S5:**
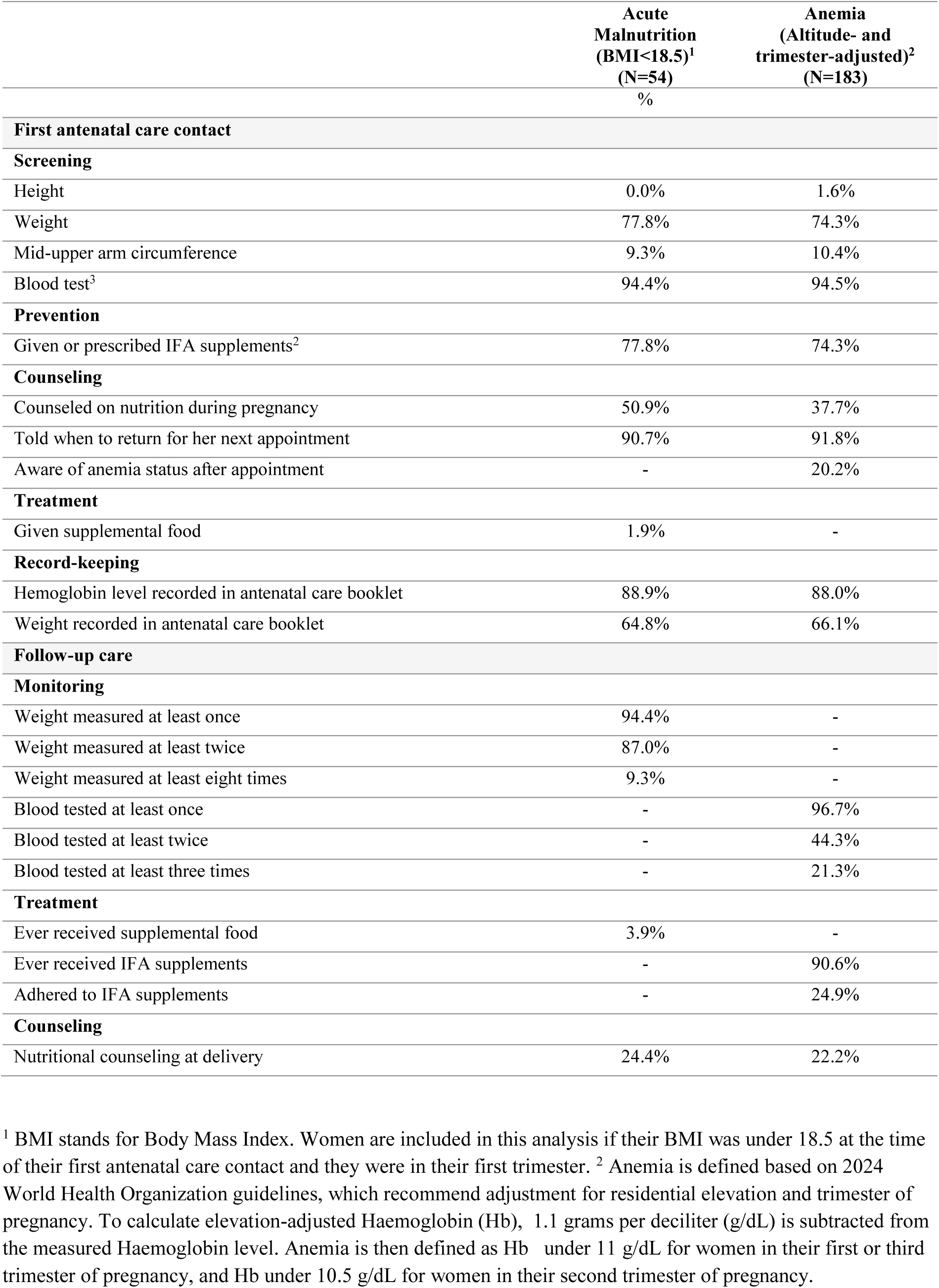
Sensitivity analysis: quality of care using alternative definitions of undernutrition.

**Figure S3:**
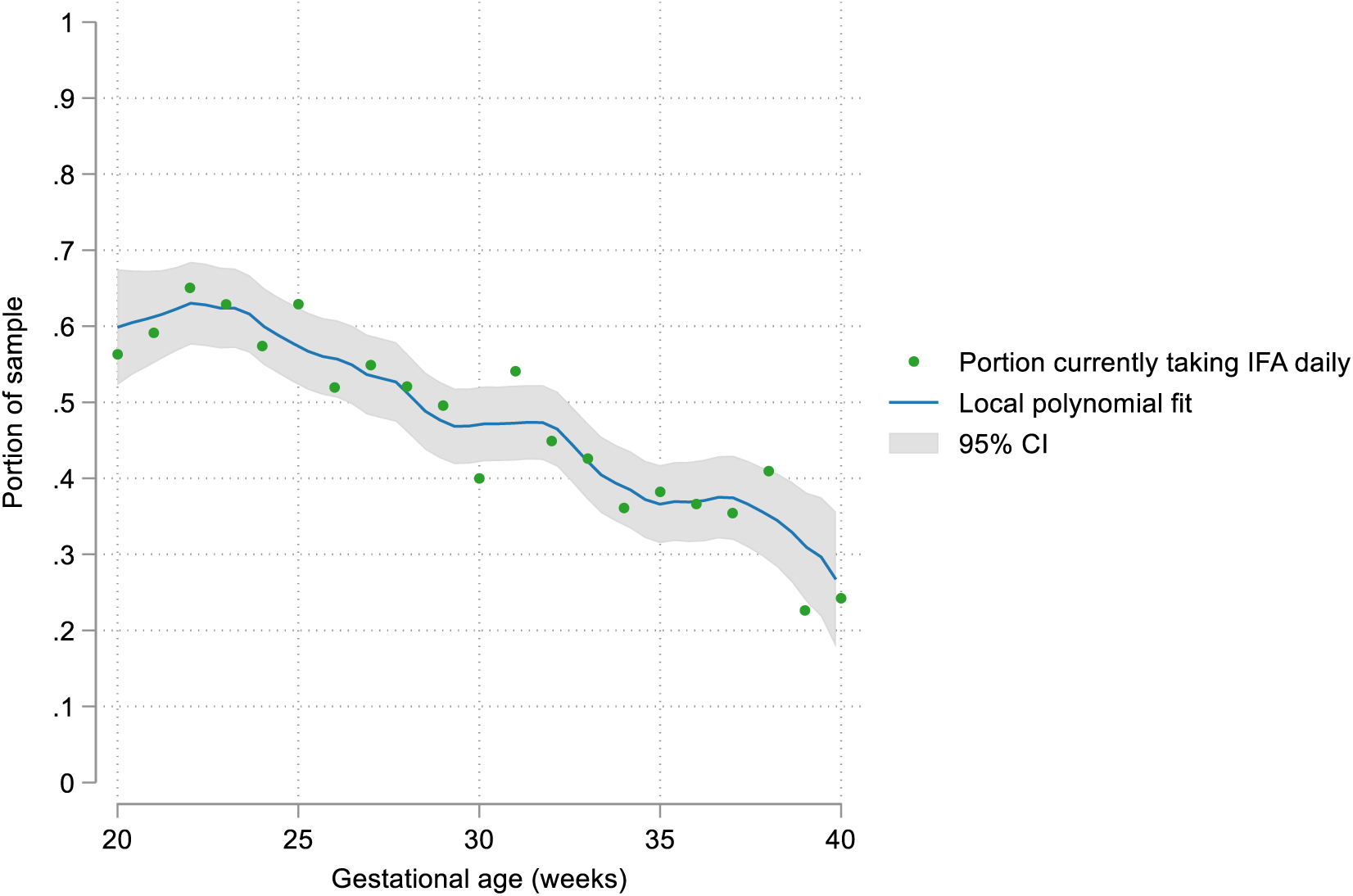
Self-reported daily adherence to iron and folic acid supplementation during pregnancy, by gestational age (restricted sample) **Notes**: This figure shows data from monthly follow-up phone interviews throughout pregnancy. The sample is restricted to N=618 women whose first antenatal care contact occurred at a gestational age of 20 weeks or earlier. Gestational age at the time of the interview is rounded to the nearest week. Green dots show the proportion of women who, during interviews at each gestational age, reported daily adherence to iron & folic acid supplementation. The blue line shows an estimated local polynomial fit, and the gray shading shows the 95% confidence interval around the line.

**Figure S4:**
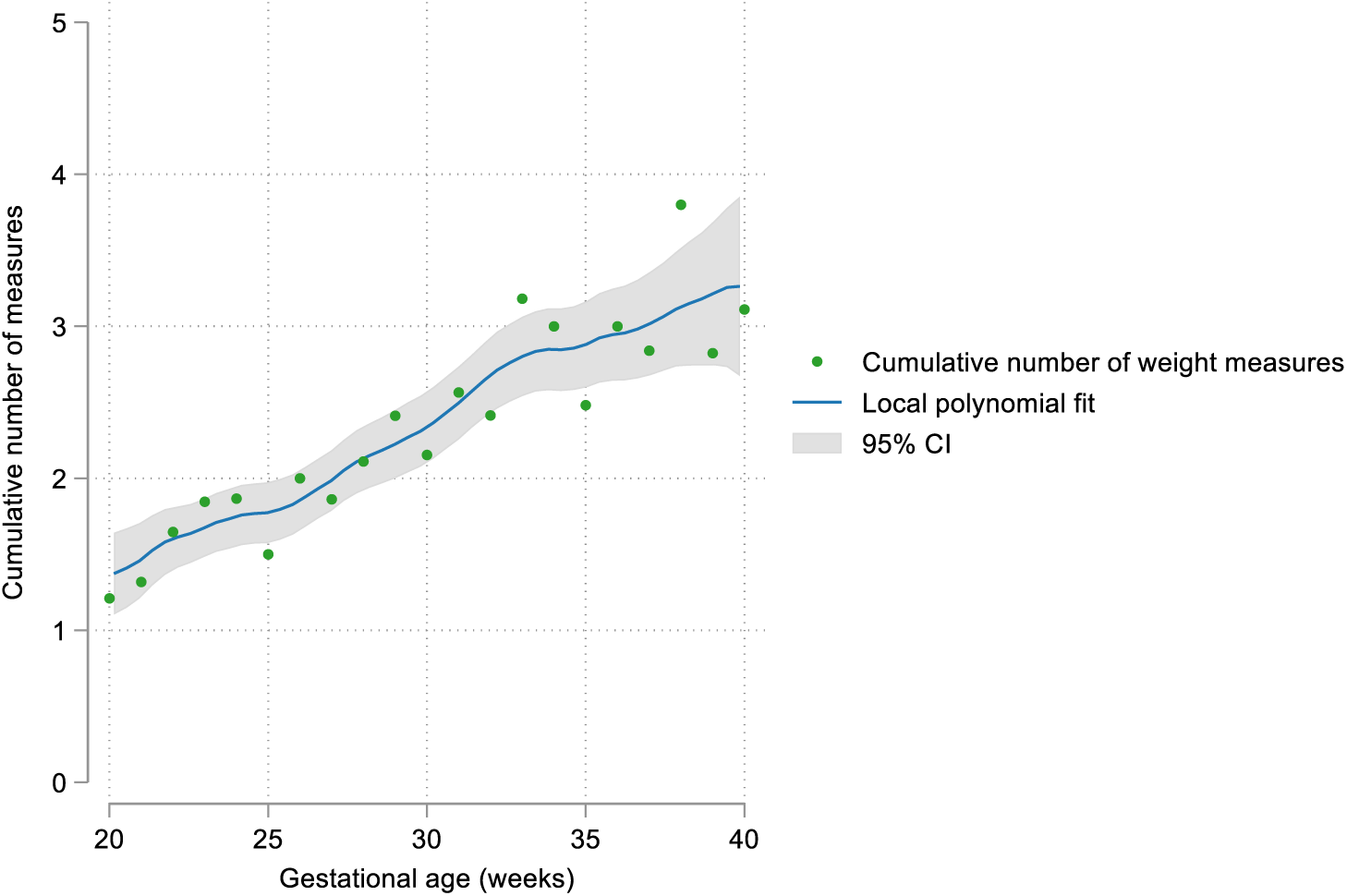
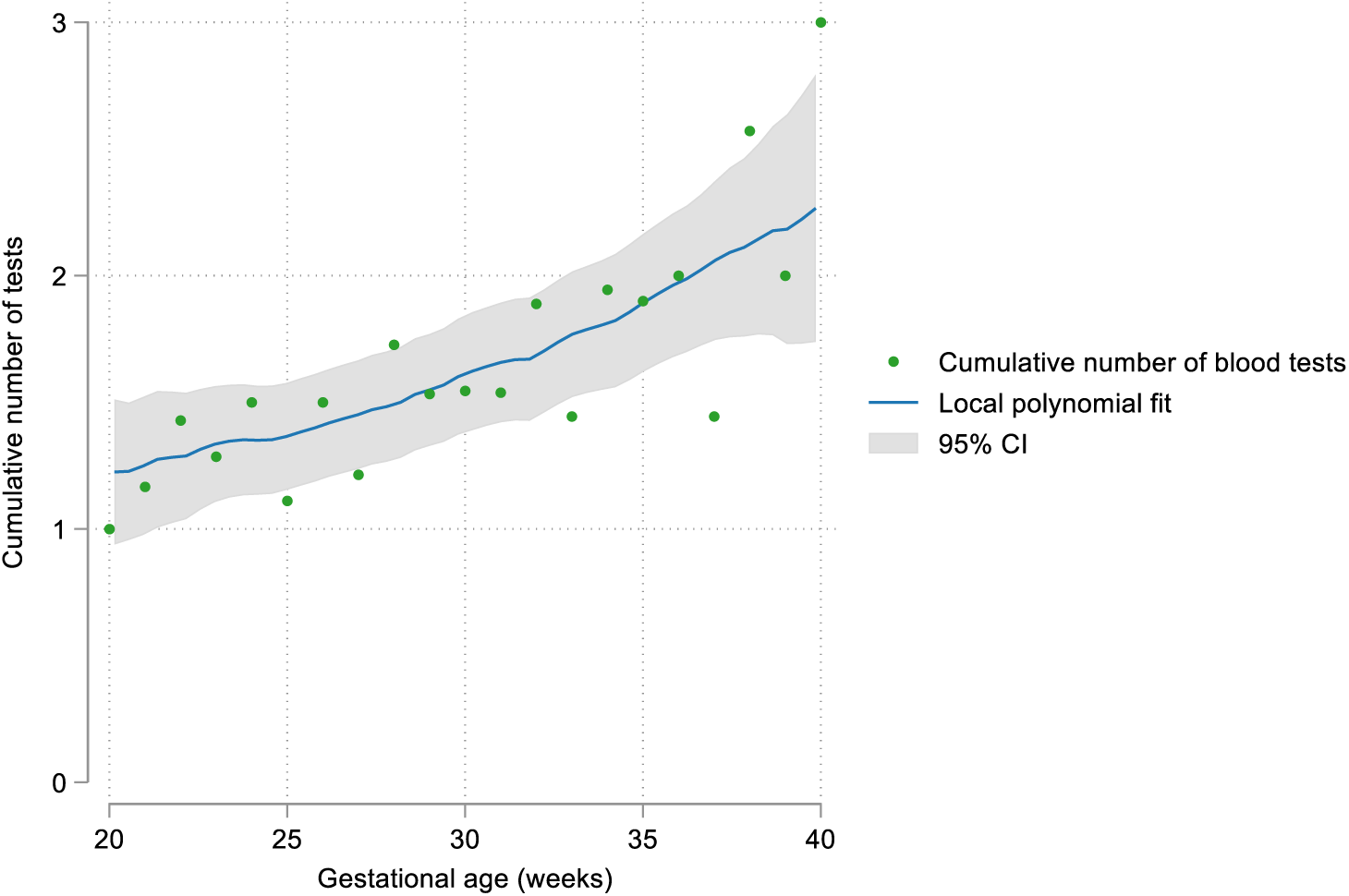
Cumulative weight monitoring and blood testing over the course of pregnancy. Panel A: Average number of cumulative weight measures by gestational age among women with acute malnutrition at the time of their first antenatal care contact (N=174) Panel B: Average number of cumulative blood tests by gestational age among women with anemia at the time of their first antenatal care contact (N=90) Note: This figure shows data from monthly follow-up interviews throughout pregnancy. Green dots show the average cumulative number of weight measures (panel A) and blood tests (panel B) by gestational age. The blue line shows an estimated local polynomial fit, and the gray shading shows the 95% confidence interval around the line. If a woman has had more than one routine ANC consultation since her last interview, and she reports that she had her weight measured (or blood tested) at least once since her last interview, then we assume that she had her weight measured (or blood tested) at all of the routine consultations since her last interview.

**Table S6:**
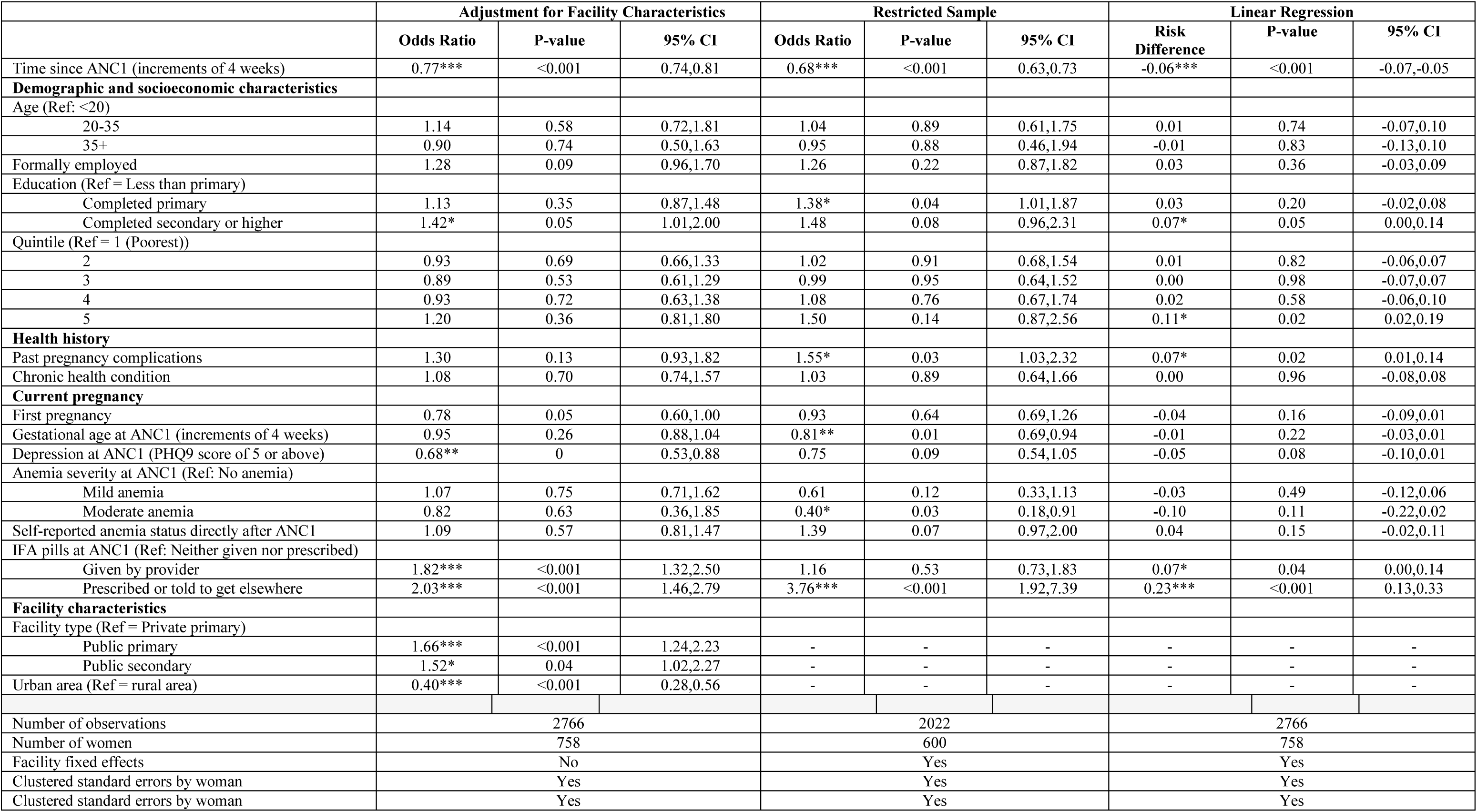
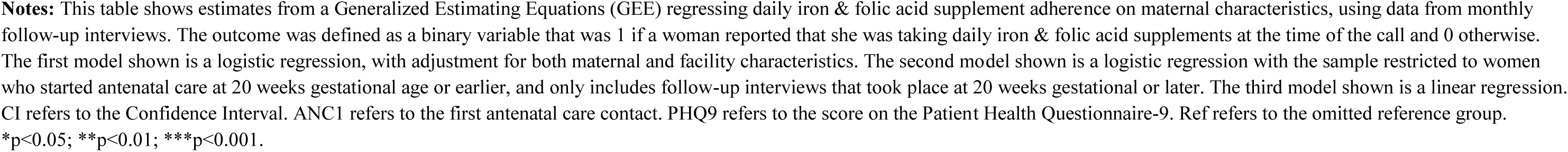
Sensitivity analyses for predictors of iron & folic acid supplement adherence.

|  | Adjustment for Facility Characteristics |  |  | Restricted Sample |  |  | Linear Regression |  |  |
| --- | --- | --- | --- | --- | --- | --- | --- | --- | --- |
|  | Odds Ratio | P-value | 95% CI | Odds Ratio | P-value | 95% CI | Risk Difference | P-value | 95% CI |
| Time since ANC1 (increments of 4 weeks) | 0.77*** | <0.001 | 0.74,0.81 | 0.68*** | <0.001 | 0.63,0.73 | -0.06*** | <0.001 | -0.07,-0.05 |
| **Demographic and socioeconomic characteristics** |  |  |  |  |  |  |  |  |  |
| Age (Ref: <20) |  |  |  |  |  |  |  |  |  |
| 20-35 | 1.14 | 0.58 | 0.72,1.81 | 1.04 | 0.89 | 0.61,1.75 | 0.01 | 0.74 | -0.07,0.10 |
| 35+ | 0.90 | 0.74 | 0.50,1.63 | 0.95 | 0.88 | 0.46,1.94 | -0.01 | 0.83 | -0.13,0.10 |
| Formally employed | 1.28 | 0.09 | 0.96,1.70 | 1.26 | 0.22 | 0.87,1.82 | 0.03 | 0.36 | -0.03,0.09 |
| Education (Ref = Less than primary) |  |  |  |  |  |  |  |  |  |
| Completed primary | 1.13 | 0.35 | 0.87,1.48 | 1.38* | 0.04 | 1.01,1.87 | 0.03 | 0.20 | -0.02,0.08 |
| Completed secondary or higher | 1.42* | 0.05 | 1.01,2.00 | 1.48 | 0.08 | 0.96,2.31 | 0.07* | 0.05 | 0.00,0.14 |
| Quintile (Ref = 1 (Poorest)) |  |  |  |  |  |  |  |  |  |
| 2 | 0.93 | 0.69 | 0.66,1.33 | 1.02 | 0.91 | 0.68,1.54 | 0.01 | 0.82 | -0.06,0.07 |
| 3 | 0.89 | 0.53 | 0.61,1.29 | 0.99 | 0.95 | 0.64,1.52 | 0.00 | 0.98 | -0.07,0.07 |
| 4 | 0.93 | 0.72 | 0.63,1.38 | 1.08 | 0.76 | 0.67,1.74 | 0.02 | 0.58 | -0.06,0.10 |
| 5 | 1.20 | 0.36 | 0.81,1.80 | 1.50 | 0.14 | 0.87,2.56 | 0.11* | 0.02 | 0.02,0.19 |
| **Health history** |  |  |  |  |  |  |  |  |  |
| Past pregnancy complications | 1.30 | 0.13 | 0.93,1.82 | 1.55* | 0.03 | 1.03,2.32 | 0.07* | 0.02 | 0.01,0.14 |
| Chronic health condition | 1.08 | 0.70 | 0.74,1.57 | 1.03 | 0.89 | 0.64,1.66 | 0.00 | 0.96 | -0.08,0.08 |
| **Current pregnancy** |  |  |  |  |  |  |  |  |  |
| First pregnancy | 0.78 | 0.05 | 0.60,1.00 | 0.93 | 0.64 | 0.69,1.26 | -0.04 | 0.16 | -0.09,0.01 |
| Gestational age at ANC1 (increments of 4 weeks) | 0.95 | 0.26 | 0.88,1.04 | 0.81** | 0.01 | 0.69,0.94 | -0.01 | 0.22 | -0.03,0.01 |
| Depression at ANC1 (PHQ9 score of 5 or above) | 0.68** | 0 | 0.53,0.88 | 0.75 | 0.09 | 0.54,1.05 | -0.05 | 0.08 | -0.10,0.01 |
| Anemia severity at ANC1 (Ref: No anemia) |  |  |  |  |  |  |  |  |  |
| Mild anemia | 1.07 | 0.75 | 0.71,1.62 | 0.61 | 0.12 | 0.33,1.13 | -0.03 | 0.49 | -0.12,0.06 |
| Moderate anemia | 0.82 | 0.63 | 0.36,1.85 | 0.40* | 0.03 | 0.18,0.91 | -0.10 | 0.11 | -0.22,0.02 |
| Self-reported anemia status directly after ANC1 | 1.09 | 0.57 | 0.81,1.47 | 1.39 | 0.07 | 0.97,2.00 | 0.04 | 0.15 | -0.02,0.11 |
| IFA pills at ANC1 (Ref: Neither given nor prescribed) |  |  |  |  |  |  |  |  |  |
| Given by provider | 1.82*** | <0.001 | 1.32,2.50 | 1.16 | 0.53 | 0.73,1.83 | 0.07* | 0.04 | 0.00,0.14 |
| Prescribed or told to get elsewhere | 2.03*** | <0.001 | 1.46,2.79 | 3.76*** | <0.001 | 1.92,7.39 | 0.23*** | <0.001 | 0.13,0.33 |
| **Facility characteristics** |  |  |  |  |  |  |  |  |  |
| Facility type (Ref = Private primary) |  |  |  |  |  |  |  |  |  |
| Public primary | 1.66*** | <0.001 | 1.24,2.23 | - | - | - | - | - | - |
| Public secondary | 1.52* | 0.04 | 1.02,2.27 | - | - | - | - | - | - |
| Urban area (Ref = rural area) | 0.40*** | <0.001 | 0.28,0.56 | - | - | - | - | - | - |
| Number of observations | 2766 |  |  | 2022 |  |  | 2766 |  |  |
| Number of women | 758 |  |  | 600 |  |  | 758 |  |  |
| Facility fixed effects | No |  |  | Yes |  |  | Yes |  |  |
| Clustered standard errors by woman | Yes |  |  | Yes |  |  | Yes |  |  |
| Clustered standard errors by woman | Yes |  |  | Yes |  |  | Yes |  |  |

**Notes:** This table shows estimates from a Generalized Estimating Equations (GEE) regressing daily iron & folic acid supplement adherence on maternal characteristics, using data from monthly follow-up interviews. The outcome was defined as a binary variable that was 1 if a woman reported that she was taking daily iron & folic acid supplements at the time of the call and 0 otherwise. The first model shown is a logistic regression, with adjustment for both maternal and facility characteristics. The second model shown is a logistic regression with the sample restricted to women who started antenatal care at 20 weeks gestational age or earlier, and only includes follow-up interviews that took place at 20 weeks gestational or later. The third model shown is a linear regression. CI refers to the Confidence Interval. ANC1 refers to the first antenatal care contact. PHQ9 refers to the score on the Patient Health Questionnaire-9. Ref refers to the omitted reference group. \*p<0.05; \*\*p<0.01; \*\*\*p<0.001.

**Figure S5:**
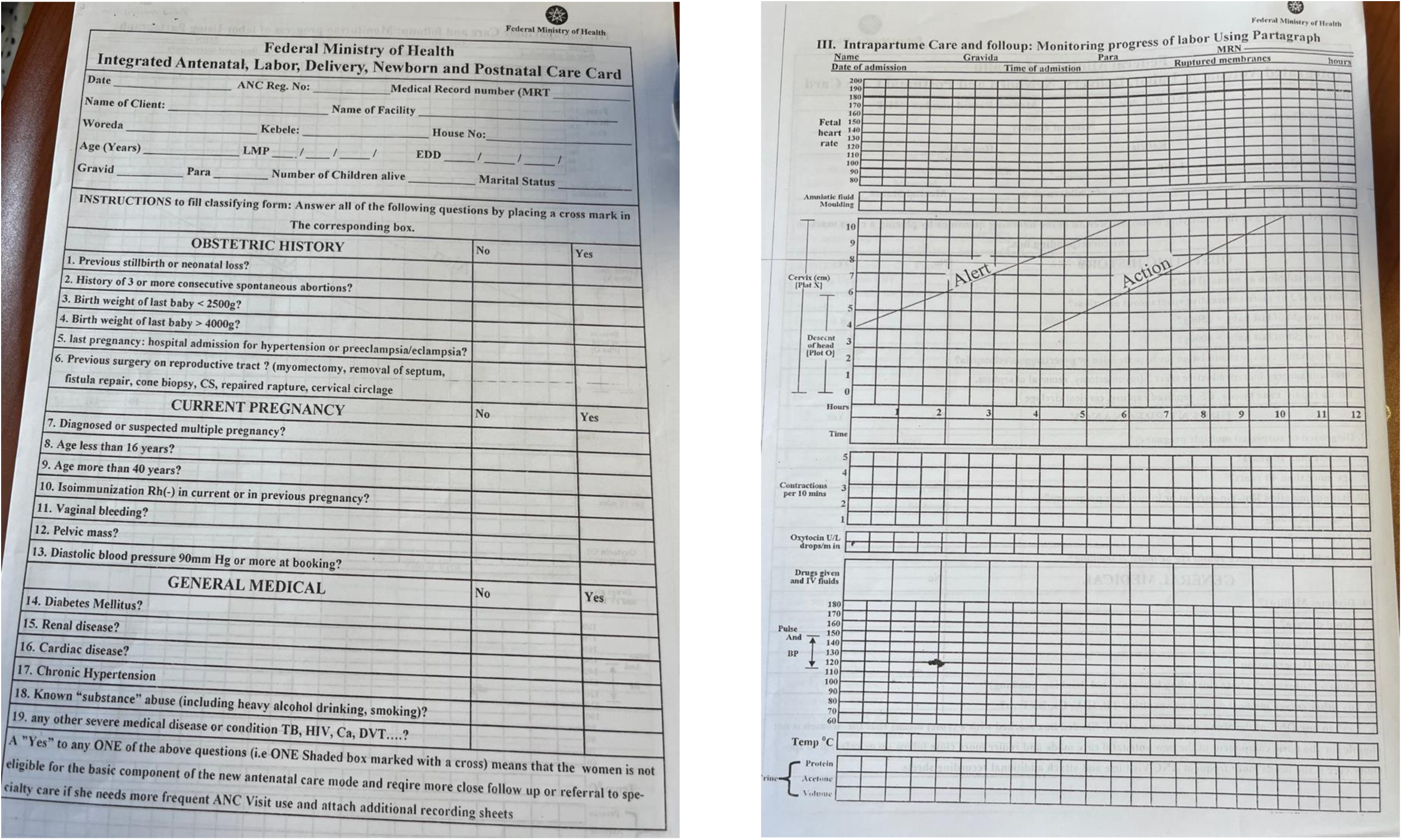
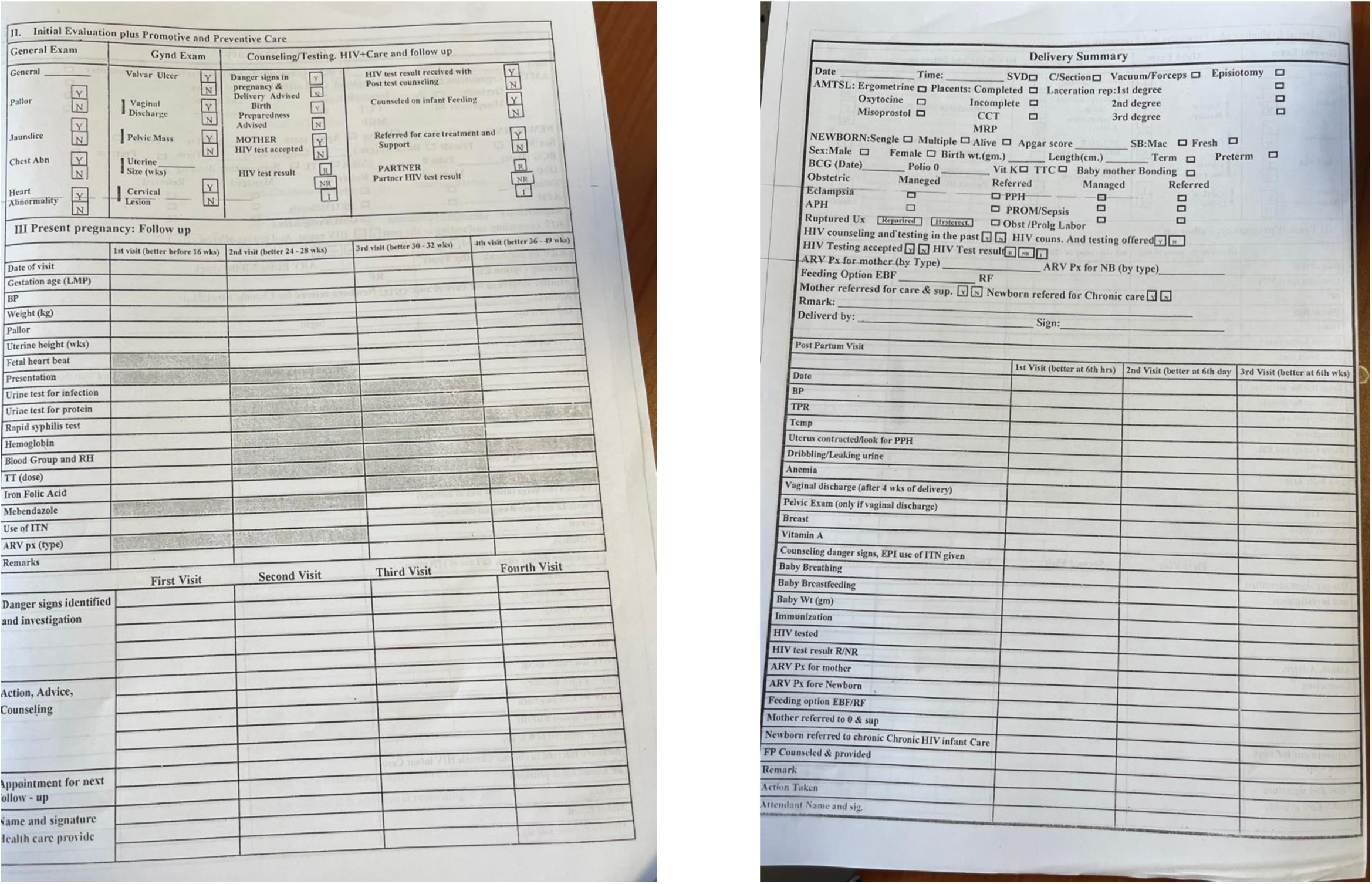
Integrated Maternal and Child Health Care Card.

## Notes

### Competing Interest Statement

The authors have declared no competing interest.

### Funding Statement

MEK received funding for the QuEST network and the eCohort study. Funding was provided by the Bill and Melinda Gates Foundation (grant number INV-005254) and the Swiss Federal Department of Foreign Affairs (grant number 81067262). The funders did not play any role in the study design, data collection and analysis, decision to publish, or preparation of the manuscript.

## References

1. Black RE, Allen LH, Bhutta ZA, Caulfield LE, De Onis M, Ezzati M, et al. Maternal and child undernutrition: global and regional exposures and health consequences. The Lancet. 2008 Jan;371(9608):243–60.

2. Black RE, Victora CG, Walker SP, Bhutta ZA, Christian P, De Onis M, et al. Maternal and child undernutrition and overweight in low-income and middle-income countries. The Lancet. 2013 Aug;382(9890):427–51.

3. Omotayo MO, Abioye AI, Kuyebi M, Eke AC. Prenatal anemia and postpartum hemorrhage risk: A systematic review and meta-analysis. J of Obstet and Gynaecol. 2021 Aug;47(8):2565–76.

4. Hofmeyr GJ, Lawrie TA, Atallah ÁN, Torloni MR. Calcium supplementation during pregnancy for preventing hypertensive disorders and related problems. Cochrane Pregnancy and Childbirth Group, editor. Cochrane Database of Systematic Reviews [Internet]. 2018 Oct 1 [cited 2024 Jun 19];2018(10). Available from: http://doi.wiley.com/10.1002/14651858.CD001059.pub5

5. Bhaskaram P. Micronutrient Malnutrition, Infection, and Immunity: an Overview. Nutrition Reviews. 2002 May 1;60(suppl_5):S40–5.

6. Keats EC, Das JK, Salam RA, Lassi ZS, Imdad A, Black RE, et al. Effective interventions to address maternal and child malnutrition: an update of the evidence. The Lancet Child & Adolescent Health. 2021 May;5(5):367–84.

7. Salam RA, Das JK, Ali A, Lassi ZS, Bhutta ZA. Maternal undernutrition and intrauterine growth restriction. Expert Review of Obstetrics & Gynecology. 2013 Nov;8(6):559–67.

8. Black MM, Walker SP, Fernald LCH, Andersen CT, DiGirolamo AM, Lu C, et al. Early childhood development coming of age: science through the life course. The Lancet. 2017 Jan;389(10064):77– 90.

9. Nandi A, Behrman JR, Kinra S, Laxminarayan R. Early-Life Nutrition Is Associated Positively with Schooling and Labor Market Outcomes and Negatively with Marriage Rates at Age 20–25 Years: Evidence from the Andhra Pradesh Children and Parents Study (APCAPS) in India. The Journal of Nutrition. 2018 Jan;148(1):140–6.

10. Victora CG, Adair L, Fall C, Hallal PC, Martorell R, Richter L, et al. Maternal and child undernutrition: consequences for adult health and human capital. The Lancet. 2008 Jan;371(9609):340–57.

11. Perumal N, Blakstad MM, Fink G, Lambiris M, Bliznashka L, Danaei G, et al. Impact of scaling up prenatal nutrition interventions on human capital outcomes in low- and middle-income countries: a modeling analysis. The American Journal of Clinical Nutrition. 2021 Nov;114(5):1708–18.

12. UNICEF. Undernourished and overlooked: a global nutrition crisis in adolescent girls and women [Internet]. New York; 2023 Mar [cited 2024 Jun 16]. (UNICEF Child Nutrition Report Series). Available from: https://www.unicef.org/media/136876/file/Full%20report%20(English).pdf

13. Ververs M tesse, Antierens A, Sackl A, Staderini N, Captier V. Which Anthropometric Indicators Identify a Pregnant Woman as Acutely Malnourished and Predict Adverse Birth Outcomes in the Humanitarian Context? PLoS Curr [Internet]. 2013 [cited 2024 Jun 13]; Available from: https://currents.plos.org/disasters/?p=7157

14. World Health Organization. WHO recommendations on antenatal care for a positive pregnancy experience. 2016.

15. Government of Ethiopia, Federal Ministry of Health. National Guideline for the Management of Acute Malnutrition in Ethiopia [Internet]. Addis Ababa; 2019 [cited 2022 May 6]. Available from: https://www.nutritioncluster.net/sites/nutritioncluster.com/files/2022-06/National%20Guideline%20for%20Management%20of%20Acute%20Malnutrition%20May%202019%20Version.pdf

16. World Health Organization. Guideline on haemoglobin cutoffs to define anaemia in individuals and populations [Internet]. 2024 [cited 2024 Jun 20]. Available from: https://iris.who.int/bitstream/handle/10665/376196/9789240088542-eng.pdf?sequence=1

17. Andersen CT, Tadesse AW, Bromage S, Fekadu H, Hemler EC, Passarelli S, et al. Anemia Etiology in Ethiopia: Assessment of Nutritional, Infectious Disease, and Other Risk Factors in a Population-Based Cross-Sectional Survey of Women, Men, and Children. The Journal of Nutrition. 2022 Feb;152(2):501–12.

18. Central Statistical Agency (CSA) [Ethiopia], ICF. Ethiopia Demographic and Health Survey 2016 [Internet]. Addis Ababa, Ethiopia, and Rockville, Maryland, USA: CSA and ICF; [cited 2024 Jun 19]. Available from: https://dhsprogram.com/pubs/pdf/FR328/FR328.pdf

19. Government of Ethiopia, Federal Ministry of Health. National Antenatal Care Guideline. Addis Ababa; 2022.

20. Abdissa Z, Alemu K, Lemma S, Berhanu D, Defar A, Getachew T, et al. Effective coverage of antenatal care services in Ethiopia: a population-based cross-sectional study. BMC Pregnancy Childbirth. 2024 Apr 27;24(1):330.

21. Gelagay AA, Belachew TB, Asmamaw DB, Bitew DA, Fentie EA, Worku AG, et al. Inadequate receipt of ANC components and associated factors among pregnant women in Northwest Ethiopia, 2020–2021: a community-based cross-sectional study. Reprod Health. 2023 May 4;20(1):69.

22. Hailu GA, Weret ZS, Adasho ZA, Eshete BM. Quality of antenatal care and associated factors in public health centers in Addis Ababa, Ethiopia, a cross-sectional study. Mordaunt DA, editor. PLoS ONE. 2022 Jun 10;17(6):e0269710.

23. Muchie KF. Quality of antenatal care services and completion of four or more antenatal care visits in Ethiopia: a finding based on a demographic and health survey. BMC Pregnancy Childbirth. 2017 Dec;17(1):300.

24. Arsenault C, Wright K, et al. The maternal and newborn health eCohort to track longitudinal care quality: survey development. Under review. 2024;

25. World Health Organization. Primary Health Care Systems (Primasys)” Case Study from Ethiopia [Internet]. 2017. Available from: https://iris.who.int/bitstream/handle/10665/341082/WHO-HIS-HSR-17.8-eng.pdf?sequence=2

26. Hanley JA. Statistical Analysis of Correlated Data Using Generalized Estimating Equations: An Orientation. American Journal of Epidemiology. 2003 Feb 15;157(4):364–75.

27. Perumal N, Wang D, Darling AM, Liu E, Wang M, Ahmed T, et al. Suboptimal gestational weight gain and neonatal outcomes in low and middle income countries: individual participant data meta-analysis. BMJ. 2023 Sep 21;e072249.

28. Negash WD, Fetene SM, Shewarega ES, Fentie EA, Asmamaw DB, Teklu RE, et al. Multilevel analysis of quality of antenatal care and associated factors among pregnant women in Ethiopia: a community based cross-sectional study. BMJ Open. 2022 Jul;12(7):e063426.

29. Government of Ethiopia. National level verified hotspot woreda classification. 2022 Jan.

30. Bahati F, Kairu-Wanyoike S, Nzioki JM. Adherence to iron and folic acid supplementation during pregnancy among postnatal mothers seeking maternal and child healthcare at Kakamega level 5 hospital in Kenya: a cross-sectional study. Wellcome Open Res. 2021 Jul 5;6:80.

31. Saragih ID, Dimog EF, Saragih IS, Lin CJ. Adherence to Iron and Folic Acid Supplementation (IFAS) intake among pregnant women: A systematic review meta-analysis. Midwifery. 2022 Jan;104:103185.

32. Temesgen H, Woyraw W, Feleke FW, Mezgebu GS, Taye K, Awoke T. Iron folic acid supplementation adherence level and its associated factors among pregnant women in Ethiopia: a multilevel complex data analysis of 2019 Ethiopian mini demographic and health survey data. Front Nutr. 2024 Feb 16;11:1348275.

33. Asmamaw DB, Debebe Negash W, Bitew DA, Belachew TB. Poor adherence to iron-folic acid supplementation and associated factors among pregnant women who had at least four antenatal care in Ethiopia. A community-based cross-sectional study. Front Nutr. 2022 Dec 8;9:1023046.

34. Desta M, Kassie B, Chanie H, Mulugeta H, Yirga T, Temesgen H, et al. Adherence of iron and folic acid supplementation and determinants among pregnant women in Ethiopia: a systematic review and meta-analysis. Reprod Health. 2019 Dec;16(1):182.

35. Ba DM, Ssentongo P, Kjerulff KH, Na M, Liu G, Gao X, et al. Adherence to Iron Supplementation in 22 Sub-Saharan African Countries and Associated Factors among Pregnant Women: A Large Population-Based Study. Current Developments in Nutrition. 2019 Dec;3(12):nzz120.

36. Birhanu Z, Chapleau GM, Ortolano SE, Mamo G, Martin SL, Dickin KL. Ethiopian women’s perspectives on antenatal care and iron-folic acid supplementation: Insights for translating global antenatal calcium guidelines into practice. Maternal & Child Nutrition. 2018 Feb;14(S1):e12424.

37. Koné S, Probst-Hensch N, Dao D, Utzinger J, Fink G. Improving Coverage of Antenatal Iron and Folic Acid Supplementation and Malaria Prophylaxis Through Targeted Information and Home Deliveries in Côte d’Ivoire: A Cluster-Randomized Controlled Trial. SSRN Journal [Internet]. 2022 [cited 2023 Feb 2]; Available from: https://www.ssrn.com/abstract=4039679

38. Abdisa DK, Jaleta DD, Tsegaye D, Jarso MH, Jaleta GD, Tolesa GF, et al. Effect of community based nutritional education on knowledge, attitude and compliance to IFA supplementation among pregnant women in rural areas of southwest Ethiopia: a quasi experimental study. BMC Public Health. 2023 Oct 5;23(1):1923.

39. Sanghvi TG, Nguyen PH, Forissier T, Ghosh S, Zafimanjaka M, Walissa T, et al. Comprehensive Approach for Improving Adherence to Prenatal Iron and Folic Acid Supplements Based on Intervention Studies in Bangladesh, Burkina Faso, Ethiopia, and India. Food Nutr Bull. 2023 Sep;44(3):183–94.

40. Lynch CD, Zhang J. The research implications of the selection of a gestational age estimation method. Paediatric Perinatal Epid. 2007 Sep;21(s2):86–96.

41. Getaneh T, Negesse A, Dessie G, Desta M, Assemie MA, Tigabu A. Predictors of malnutrition among pregnant women in Ethiopia: A systematic review and meta-analysis. Human Nutrition & Metabolism. 2021 Dec;26:200131.

42. Zewude SB, Beshah MH, Ahunie MA, Arega DT, Addisu D. Undernutrition and associated factors among pregnant women in Ethiopia. A systematic review and meta-analysis. Front Nutr. 2024 May 6;11:1347851.

43. Geta TG, Gebremedhin S, Omigbodun AO. Prevalence and predictors of anemia among pregnant women in Ethiopia: Systematic review and meta-analysis. Kupfer G, editor. PLoS ONE. 2022 Jul 27;17(7):e0267005.

44. Kennedy E, Mersha GA, Biadgilign S, Tessema M, Zerfu D, Gizaw R, et al. Nutrition Policy and Governance in Ethiopia: What Difference Does 5 Years Make? Food Nutr Bull. 2020 Dec;41(4):494–502.

